# Enhanced Detection and Genotyping of Disease-Associated Tandem Repeats Using HMMSTR and Targeted Long-Read Sequencing

**DOI:** 10.1101/2024.05.01.24306681

**Authors:** Kinsey Van Deynze, Camille Mumm, Connor J. Maltby, Jessica A. Switzenberg, Peter K. Todd, Alan P. Boyle

## Abstract

Tandem repeat sequences comprise approximately 8% of the human genome and are linked to more than 50 neurodegenerative disorders. Accurate characterization of disease-associated repeat loci remains resource intensive and often lacks high resolution genotype calls. We introduce a multiplexed, targeted nanopore sequencing panel and HMMSTR, a sequence-based tandem repeat copy number caller. HMMSTR outperforms current signal- and sequence-based callers relative to two assemblies and we show it performs with high accuracy in heterozygous regions and at low read coverage. The flexible panel allows us to capture disease associated regions at an average coverage of >150x. Using these tools, we successfully characterize known or suspected repeat expansions in patient derived samples. In these samples we also identify unexpected expanded alleles at tandem repeat loci not previously associated with the underlying diagnosis. This genotyping approach for tandem repeat expansions is scalable, simple, flexible, and accurate, offering significant potential for diagnostic applications and investigation of expansion co-occurrence in neurodegenerative disorders.

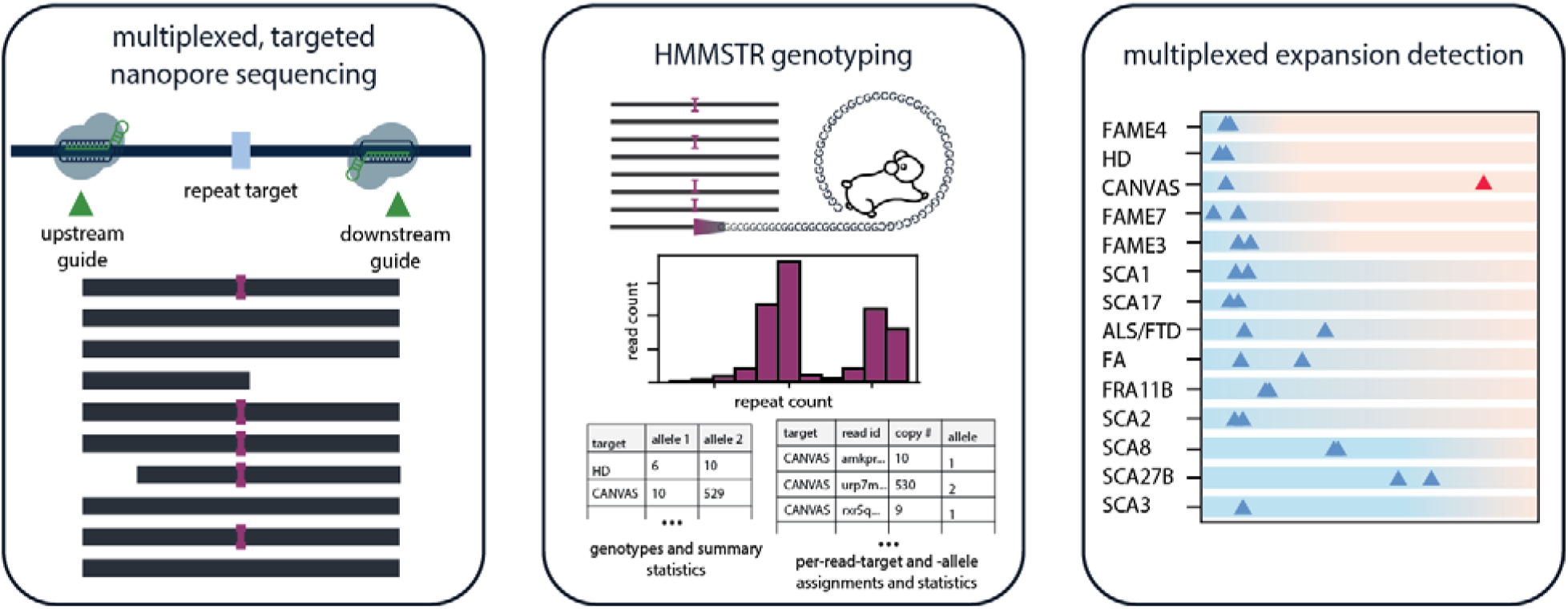

## Introduction

Tandem repeat (TR) sequences constitute about 8%(1) of the human genome, and more than 50 neurodegenerative conditions are associated with short tandem repeat (STR) expansions. Some examples of tandem repeat expansion disorders include amyotrophic lateral sclerosis (ALS) and frontotemporal dementia (FTD), polyglutamine-associated spinocerebellar ataxias, Huntington’s disease, and myotonic dystrophy(2). A number of these neurodegenerative conditions exhibit overlapping clinical symptoms. Thus, when screening for pathogenic expansions, multiple regions need to be examined. Current molecular methods for tandem repeat sizing, such as repeat primed PCR and southern blotting, genotype only one locus at a time and are labor intensive. As such, current diagnostic methods remain inefficient (3).

Alternatives to these methods include high-throughput sequencing approaches. The application of short-read sequencing techniques to tandem repeat genotyping is limited, as the number of repeat motif copies needed for pathogenicity often exceeds the length of a single read. Thus their ability to characterize large, low complexity regions has stifled their adoption in clinical settings(4–8). Long-read sequencing methods, such as PacBio and Oxford Nanopore Technologies (ONT), provide an attractive alternative that can sequence through disease associated regions despite their length and complex motif structure. However, these methods come with their own challenges, including relatively high cost and decreased accuracy in repetitive regions(9–11).

While whole-genome, long-read sequencing has been revolutionary in characterizing tandem repeats, it remains resource intensive. Compared to PacBio, ONT sequencing benefits from being more cost effective and accessible. Additionally, multiple targeted sequencing methods have been developed to provide efficient characterization of genes and regions of interest. Targeted sequencing, or the ability to selectively sequence regions of interest, offers an advantage over whole genome sequencing (WGS) by decreasing the amount of sequencing needed to obtain high coverage over targets. Thus, WGS can require more reagents, input, and time to obtain the coverage necessary for genotyping at repetitive regions of interest (12).

Recently, two techniques have been developed for targeted sequencing using ONT. The first, ReadFish, uses computational tools to select for target fragments during a live sequencing experiment (12). In 2022, this technique was used to successfully genotype disease-associated tandem repeats using the ReadFish API. The second targeted sequencing approach, termed nanopore Cas9 Targeted sequencing (nCATs), uses CRISPR-Cas9 to selectively sequence regions of interest using RNA guides and was found to outperform computational enrichment for tandem repeat genotyping (9, 13–16). However, the application of nCATS to comprehensively genotyping disease-associated tandem repeats has yet to be extended beyond common ataxias found in European populations (15).

Targeted sequencing techniques have been used in conjunction with existing long-read sequencing bioinformatic genotyping tools and significantly improve on WGS for tandem repeat genotyping. Various bioinformatics tools have been developed to overcome sequencing errors for copy number determination from ONT long-read sequencing data. However, many are designed for WGS and prioritize the discovery of novel, large expansions rather than accurately genotyping specified targets (17–19).

Methods designed for PacBio HiFi data, such as TRGT(20), rely on the generation of consensus sequences from highly accurate circular consensus sequencing (CCS). However, comparatively high error rates in repetitive regions, which can impede accurate tandem repeat genotyping, necessitate accounting for errors in ONT reads (9). Thus, nanopore-specific, signal-based methods have emerged as a promising approach for directly calling tandem repeat copy numbers from nanopore signal data to minimize errors introduced by basecalling (9, 13, 14). These methods have demonstrated success with targeted sequencing (14, 15, 21). However, they suffer both runtime and storage capacity burdens due to the need for storing and processing the signal data (9).

In response to the current limitations and challenges of genotyping the wide range of disease-associated tandem repeats in a cost-effective manner, our study aims to establish a scalable and easily extendable sequencing and bioinformatics workflow for accurate and efficient copy number determination capable of interrogating all known disease associated repeat expansion loci. Our strategy combines a multiplexed nCATS approach using a guide pool to target over 50 disease-associated repeat expansion loci on a single ONT MinION flow cell, the most comprehensive panel to date (15). Alongside this, we introduce a profile Hidden-Markov Model STR (HMMSTR) copy-number caller optimized for sequence-based targeted sequencing data. HMMSTR models ONT errors and aims to combine the workflow and accuracy of signal-based copy number callers with the efficiency of sequence-based methods.

## Materials and Methods

### Plasmid constructs

Four sets of plasmids were constructed using a pcDNA3.1 backbone that contained distinct motifs (AAAAG, AAGGG, GGGGC, and CGG) and two to five motif copy numbers (Table S1). The plasmids were restriction enzyme digested, pooled based on motif, and sequenced as described in Mumm et al (22) using ONT R9.4.1 Flongle flow cells and SQK-LSK110 kit. Next, the data was basecalled with Guppy 5.0.13. Table S1 lists the specific motifs, copy number and restriction enzymes used for the digestion reactions for each plasmid sequenced. HMMSTR was run on all constructs using 200bp flanking sequence from each backbone, the expected number of peaks as max_peaks parameter, and either Gaussian mixture modeling (GMM) or kernel density estimation (KDE) as the preferred peak-calling method based on the noise level of each construct (eg KDE was used for CGG plasmid construct).

### The HMMSTR model

The HMMSTR model is a modified version of a profile HMM (23). The model is made up of 5 distinct sections: the upstream genome state, prefix states, repeat motif states, suffix states, and downstream genome state (Figure 1A). The prefix, repeat, and suffix states follow a three layer (match, insertion, deletion) profile HMM structure. Match state emission probabilities are encoded with the expected base at each position in either the flanking sequence or the repeat motif while insertion state emissions follow a uniform distribution across all observed bases and the deletion states encode a silent character. Emission probabilities at match states reflect mismatch rates based on the expected base at a given position and transition probabilities encode expected rates of insertions and deletions. One copy of the expected motif is encoded in the model and edges are added between the last states in the motif to states at the beginning of the repeat section (Figure 1A). This allows the Viterbi algorithm to find paths through as many repeat motifs as are found in the given sequence. Our model allows for local alignment to the tandem repeat and the direct flanking sequence through the use of genome states with emission probabilities following a uniform distribution (default).

**Figure 1.**
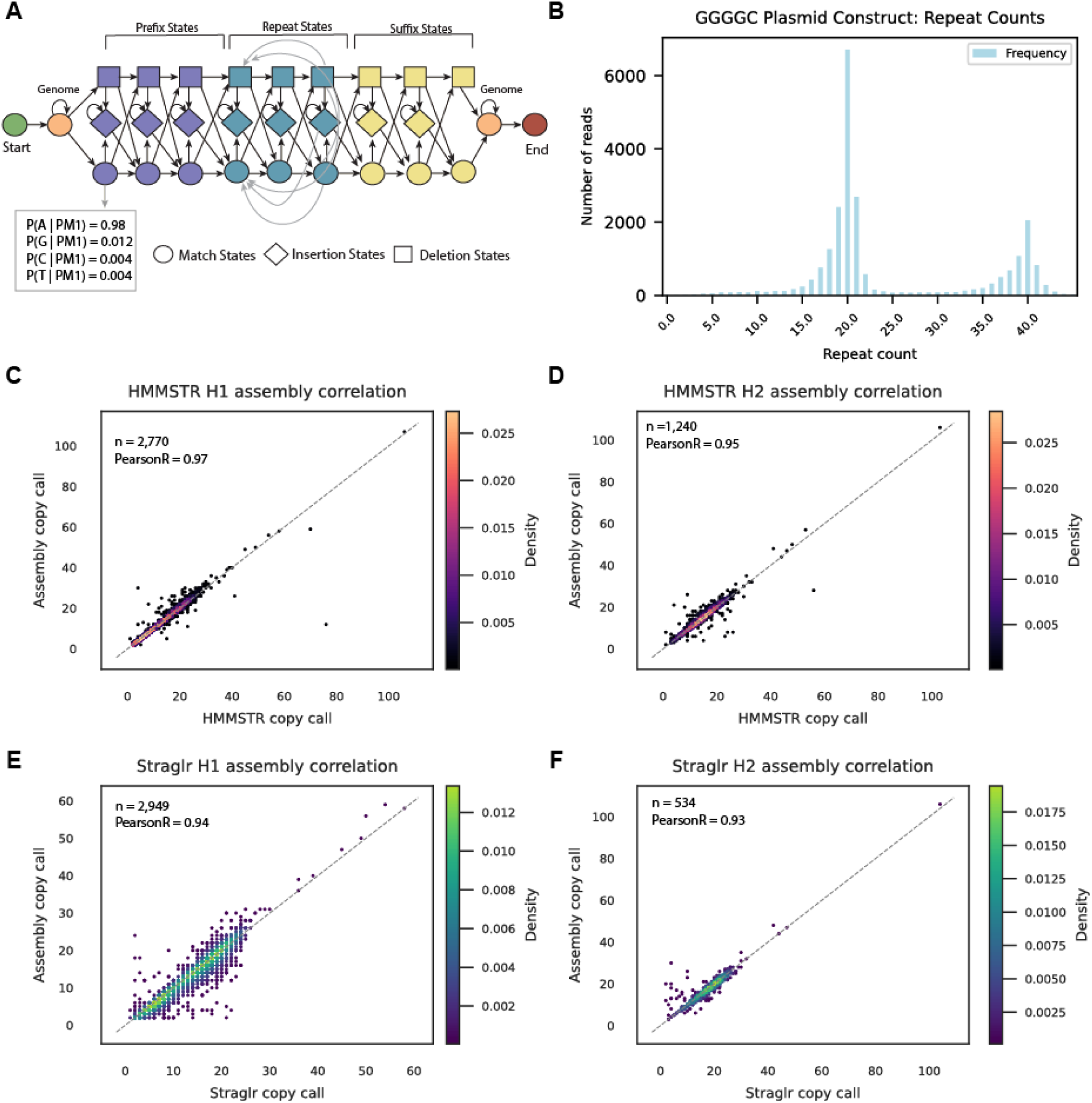
HMMSTR concordance with ground truth sets. (**A**) A breakdown of the HMMSTR model including sample emission probabilities for a position with an expected ‘A’ nucleotide. (**B**) Results from GGGGC plasmid benchmarking construct with target repeat lengths 21 and 41, HMMSTR calls 20 and 40. (**C**) Correlation between HMMSTR copy number calls from GM12878 ONT dataset and PacBio HiFi assembly copy numbers estimated by Tandem Repeat Finder (TRF) (27) on GM12878. (left) Correlation between homozygous loci and largest allele calls (n=2770 loci) and (right) correlation between smallest allele calls (n=1240). (**D**) Correlation between Straglr copy number calls from the same ONT dataset and assembly copy number estimates. (left) Correlation between homozygous loci and largest allele calls (n=2949 loci) and (right) correlation between smallest allele calls (n=534).

### Model parameter estimation

Baum-Welch was run on sequences obtained by the alignment of plasmid sets to plasmid backbone sequence per expected repeat count to estimate model parameters (24). While Baum-Welch failed to converge, multiple parameter estimates at later iterations were stable and corresponded well with literature on the same sequencing chemistry for all emission probabilities and combined deletion-insertion rate(10). Notably, Baum-Welch successfully recovered previously reported bias in substitution errors. All other parameters were estimated based on literature. Model parameters can be updated as chemistries and basecallers improve or for custom use (see HMMSTR documentation: https://github.com/Boyle-Lab/HMMSTR).

### Modification of the Viterbi algorithm

The Viterbi (25) algorithm was modified to allow for paths through deletion states without requiring labeled deletions in the observed sequence prior to the run (i.e. pre-alignment to a perfect sequence). We added a third dynamic programming trellis to keep track of when a transition through a deletion state was optimal and allowed for horizontal paths through the traceback trellis to account for these transitions. In this way, we can determine when to index horizontally instead of diagonally and add a deletion character as opposed to an observed character from the input sequence.

### The HMMSTR workflow

#### Read processing

Input flanking sequences are aligned to each read in the sample independently using Mappy (version 2.24) (26). A given read is considered for downstream analysis if (1) it has a valid alignment to both the prefix and suffix of at least one target, (2) the prefix and suffix for at least one target are in the correct orientation with respect to the strand and (3) the prefix and suffix mapq scores exceed the score cutoff threshold (default: mapq 30). If the read satisfies these requirements, it is assigned to all qualifying targets (multiple targets may be considered).

Once a read passes the initial filtering and assignment step, its sequence is subsetted to include 400bp flanking the prefix and suffix alignment positions. If this exceeds the length of the read, this subset is decreased accordingly. This step is used to decrease the runtime of the Viterbi step (which scales to observation length linearly). We choose to keep the larger flanking sequence to mitigate the effect of poor alignment and ensure use of the entire directly-flanking sequence.

#### Repeat motif counting

Repeat copy numbers were counted by taking the total length of the identified repetitive region in the labeled sequence, subtracting the number of insertions identified, and dividing by the length of the given motif. Note that deletions are accounted for in the labeled sequence and are thus included in this calculation.

#### Identifying and filtering outlier repeat copy numbers

HMMSTR has an option to filter outlier repeat counts. This can be useful when dealing with larger datasets where there are more likely off-target reads, high coverage datasets with large tails, or PCR products with amplification bias. We designate outliers using the interquartile range (IQR) of the repeat copy number data for a given target. Reads with copy numbers that are outside of this range will be filtered before peak calling is performed.

#### Summary statistics and peak calling

By default, HMMSTR chooses between Kernel Density Estimation (KDE) with a gaussian kernel and Gaussian Mixture Modeling (GMM) for calling genotypes from the per-read copy number data. Both methods have advantages in distinct situations depending on the distribution of the data.

KDE resolves homozygotes better than GMM in situations where the data has a narrow IQR with few outliers. In this situation, the GMM will overcall heterozygous regions while the KDE is more able to distinguish hetero- and homozygosity. However, the GMM can more accurately detect distributions with larger distance between means and is less often skewed by outliers. A third case is also considered where the quantile range of the data is narrow but there exists few outliers. In this case, a KDE is optimal with the exception of the outliers. Thus, in this case outliers are filtered and a KDE is used to call the genotype since the majority of the data remains in the IQR. This choice can also be overridden as an input along with multiple KDE parameters.

By default, HMMSTR assumes a diploid sample and the maximum number of alleles called is set to 2, however this is a customizable parameter in HMMSTR.

#### Preprocessing of WGS benchmarking sets

Both the GM12878 ONT WGS dataset (https://github.com/nanopore-wgs-consortium/NA12878/blob/master/Genome.md) and PacBio CCS WGS dataset (https://www.ncbi.nlm.nih.gov/bioproject/PRJNA540705) were downloaded and subset by chromosome then target regions per chromosome using samtools. All subset bam files were then converted to fasta format for HMMSTR input using samtools fasta.

### WGS benchmarking pipeline

#### Loci selection

From our original benchmarking set, we filtered all loci annotated with less than 80% motif composition or reported multiple possible repeat motifs or overlapping annotations for the same locus according to the hg38 simple repeats track (27). This resulted in 11,035 regions. We used the PacBio HiFi GM12878 assembly from the Human Genome Structural Variation Consortium (HGSVC) (https://ftp.1000genomes.ebi.ac.uk/vol1/ftp/data_collections/HGSVC2/working/20200417_Marsc <u>hall-Eichler_NBT_hap-assm/</u>). Assembly genotypes were found by aligning the 200bp flanking sequence to each assembly haplotype followed by copy number determination using TandemRepeatFinder (TRF) (27).

For the heterozygous benchmarking, we started with all loci in the simple repeats track and filtered in the same manner as the initial benchmark. We then extracted the 2000bp flanking sequence from hg38 and aligned these sequences to the PacBio HiFi assembly (28). From the loci that had valid alignments both upstream and downstream of a given tandem repeat, the number of repeat copies was determined by running TRF. All regions called as homozygous by TRF were filtered resulting in 122,864 heterozygous regions. We further filtered all heterozygous regions where the 200bp flanking sequences were completely masked in the reference genome as well as regions where the 30bp directly flanking the repeat was repeated within 200bp of the repeat. The latter was applied to reduce ambiguity in which prefix or suffix sequence is chosen by HMMSTR when using a model with a 100bp flanking sequence, however these regions may still be accurately resolved with larger models. This resulted in 19,256 target regions.

#### HMMSTR and Straglr benchmarks

HMMSTR was run with a 90 base pair flanking sequence model for the original GM12878 benchmarking and 100 base pair flanking sequence model for the heterozygous benchmarking. Default settings were used with the exception of the discard outliers option and the PacBio alignment flag (for the PacBio runs only) on each dataset separately. Straglr was run on the same regions with default settings. We filtered homopolymers, regions with less than two read coverage, and regions with 200bp of masked sequence flanking the repeat of interest according to the repeatmasker (http://www.repeatmasker.org). Both the full datasets used and the regions of interest had approximately 30x coverage and the number of regions genotyped by each tool has been reported in Supplementary Figure 13 for both GM12878 benchmarks.

#### Assigning haplotype sets

The copy number calls were sorted by size and labeled as heterozygous and homozygous. In order to conduct a fair comparison to the assembly and assess the specificity and sensitivity of each tool in sizing heterozygous alleles, we sorted and assigned calls to two sets as follows: allele 1 (H1) contains the set of copy number calls where both the assembly and tool calls were homozygous or both heterozygous (larger allele). In the case where the assembly and tool call disagreed with homozygous/heterozygous call, we assigned a heterozygous tool call to H1 if the homozygous assembly call was closer to the larger allele call and vice-versa. The allele 2 (H2) set was constructed as the complement of H1 such that the smaller allele for all regions that were called heterozygous by both the assembly and the given tool were included and all regions where the tool called a heterozygote with the assembly call closer to the smaller tool call.

#### Assessing correlation

PearsonR correlation was calculated for each of the sets with the following filters: all regions had a valid copy number call from both HMMSTR and Straglr; all regions had at least 2 supporting reads in the given dataset; and the 200bp flanking regions (on both sides) could not be entirely masked by repeatmasker.

#### Upset plots

Sets used to construct upset plots were created by separating regions called heterozygous in one, two, or all datasets (ONT, PacBio, and Assembly) for both Straglr and HMMSTR separately. All regions called by a given tool were included in this analysis regardless of if a given region was called by the other tool.

### CHM13 benchmarking

HMMSTR and Straglr were run on regions defined by Fang et al.(9). This set was 439 regions over 200bp not within 500bp of another STR. Basecalled reads (Guppy 5.0.7) were downloaded from the Telomere-to-telomere consortium github at https://github.com/marbl/CHM13. Reads were aligned to the CHM13 (v2.0) using minimap2 (version 2.26). Reads were then extracted from regions of interest using samtools. HMMSTR was run with discarded outliers, a mapq filter of 60, and maximum peaks parameter of 1. Straglr was run with maximum peaks of 1 and default settings.

Downsampling analysis was performed by running samtools view -s with fractions of 0.5, 0.33, 0.15, 0.10, and 0.05 for each chromosome. HMMSTR was then run with the parameters stated above.

### Defining disease-associated regions

The table outlining the known repeat expansions underlying neurological disorders was created through the adaptation of multiple literature sources combined with manual curation from publicly available genomic data found at NCBI (2, 29–33).

### Defining normal, intermediate and pathogenic ranges

Normal, intermediate, and pathogenic repeat copy number ranges for each disease-associated loci were defined according to Chaisson et al. (33) or Stripy (32). The intermediate range was defined as any copy number between the upper limit of the normal range and the lower limit of the pathogenic range.

### Swimlane plot genotype calls

Disease-associated regions were genotyped with HMMSTR with mapq cutoff of 60 and default parameters with the exception of few loci which required 200bp flanking sequence for optimal target specificity (see github). The flanking filter flag was passed to discard reads with spurious sequence.

### Estimating softclip or non-through read repeat copy numbers with HMMSTR

Softclip reads were identified using samtools view on sample bam files using samtools view. Reads spanning the entirety of the repeat region per target were filtered based on HMMSTR output such that only softclip reads were left. The softclip reads per target were then converted to fasta format independently. Softclip reads were kept separate per target to ensure correct target assignment in the absence of adequate flanking sequence on both sides of the given repeat. Softclip read repeat copy numbers were found with HMMSTR with the following parameters:

–flanking_size 60

–mode sr

–mapq_cutoff 0

–use_full_read

where the prefix and suffix were made up of the concatenation of the respective repeat motif. This circumvented the need for the read to contain any flanking sequence not included in the softclip reads. These parameters allow mappy to align the short (60bp) flanking sequence inputs to the reads successfully and ensure the full read is considered regardless of which repeat the input flanking sequence aligns to. The mapq cutoff ensures the reads will not be thrown out due to multimapping of the repetitive prefix and suffix inputs. The number of repeats found in the flanking sequence were added to the total repeat copy number reported by HMMSTR.

### Calculating motif composition

uTR(34) was run on reads from each allele per target independently. Motif decompositions were then processed using a custom script, and composite motif compositions were constructed per individual allele as follows: the number of occurrences of a given motif per read was calculated; the median number of occurrences of each motif was taken across all reads; the final composition was calculated as the percent of the total median motif occurrences a given motif made up.

### Cell culture

The GM12878 cell line was obtained from the NIGMS Human Genetic Cell Repository at the Coriell Institute for Medical Research. GM12878 was cultured at 37 °C, 5% CO_2_ in RPMI 1640 media with L-glutamine (11875093, ThermoFisher), and supplemented with 15% fetal bovine serum (10437028, ThermoFisher) and 1X antimycotic–antibiotic (15240112, ThermoFisher). Cells were regularly passed and the media replenished every 3 days.

Dermal fibroblasts were obtained from consenting patients clinically diagnosed with CANVAS spectrum disorder and genetically confirmed to possess biallelic *RFC1* expansions and iPSCs were cultured as described in Maltby et al.(35). ALS patient fibroblasts originated from a 66 yo male Michigan Medicine patient and obtained as skin biopsy punch under local IRB approval (HUM00030934). Tissues were obtained from UV protected areas such as behind the knee/back of the upper leg using a 5.0 mm biopsy punch. The biopsy punch was cut to remove subcutaneous fat, and then pulverized and plated in a small dish containing Fibroblast Medium (DMEM, 10% (v/v) FBS, 1% (v/v) 100x NEAA, 1% (v/v) Pen/Strep, 1.5% (v/v) 1M HEPES) at 37 °C and 5% CO2 until fibroblasts emerged from the tissue and were maintained at low passage number prior to expanding and banking.

Six post-mortem cerebellum samples were obtained from the University of Michigan Brain Bank with informed consent of the patients or their relatives and the approval of the local institutional review boards (IRB).

### Genomic DNA preparation

High molecular weight genomic DNA (HMW gDNA) was extracted from CANVAS patient derived iPSC cell pellets (∼30M cells) using the salting out method detailed in McDonald et al. ALS/FTD fibroblast and GM12878 LCL HMW gDNA was extracted using the Monarch® HMW DNA Extraction Kit for Cells and Blood (T3050L, NEB) following the manufacturers’ instructions. Brain tissue HMW gDNA was extracted from 50mg sections using the Monarch® HMW DNA Tissue Extraction Kit (T3060L, NEB) following the manufacturer’s protocol with the following changes in the lysis step. 40μL of 10 mg/mL Proteinase K (3115879001, Roche) was added to 580μL of Tissue Lysis Buffer. The tissue was placed at 56°C for 15 minutes on a ThermoMixer (Eppendorf) with 2000 rpm mixing then incubated at 56°C for 30 minutes without agitation.

### Guide selection

Guide RNAs (sgRNAs) were designed according to ONT’s best practices for nCATS (https://community.nanoporetech.com/docs/plan/best_practice/targeted-amplification-free-dna-sequencing-using-crispr-cas/v/eci_s1014_v1_revf_11dec2018). In Panel 1, three guides were designed 2-5kb upstream and downstream of 54 targets. These 20bp sgRNAs guides were chosen using a pipeline consisting of command line ChopChop and CRISPRon (36, 37). The nanopore enrichment model was used with ChopChop, where we specified hg38 regions 2-5kb upstream and downstream of the disease-associated tandem repeat target. The candidate guides were then filtered based on strand for nCATs directionality and further scored using an in-house adaption of CRISPRon.

We then inserted the 20bp sgRNA guide into the following template for pooled amplification and transcription based on Gilpatrick et al 2022 (38).

5’-TAATACGACTCACTATAG**-*20nt-seq\***

**-** GTTTTAGAGCTAGAAATAGCAAGTTAAAATAAGGCTAGTCCGTTATCAACTTGAAAAAGTGGCACCGAGTCGGTGCTTTT

In subsequent panel iterations (Panel 2 and 3), guides were added for additional targets and removed for high off-targeting (Tables S2-7).

### Guide preparation

The oligo pool from TWIST Biosciences was resuspended to 1ng/μL and 1ng was used for amplification with the primers below according to manufacturer instructions with the following PCR conditions: PCR conditions: 95°C 3 min, 20 cycles of 98°C 20 sec, 65°C 15 sec, and 72°C 15 sec, 72°C 10 min and then held at 12°C.

T7 anchored fwd (5’-CGCGCGTAATACGACTCACTATAG-3’)

T7 rev (5’-AAGCACCGACTCGGTGCC-3’)

The product of this reaction (24μL) was then used as input for an 8x reaction in a second round of amplification in an 8 tube strip with the same PCR conditions.

The final amplification product was pooled from the tube strip and cleaned up using the QIAquick PCR Purification kit (28104, Qiagen) and 500ng was used for *in vitro* transcription using the NEB HiScribe T7 RNA synthesis kit (E2040S, NEB) according to kit instructions. The sgRNA was purified using a Trizol/chloroform clean-up followed by ethanol precipitation, as detailed in McDonald et al.

### Library preparation

nCATS library preparation was performed following McDonald et al (39) with adaptations for LSK114 and pore R10.4.1 chemistry. First 7.5ug (or 30ul of gDNA at a concentration greater than 100ng/ul) was dephosphorylated in a 40μL reaction with 6μL Quick CIP (M0525S, NEB) and 4μL 10X rCutSmart buffer (B7204S, NEB). This reaction was inverted and gently tapped to mix and then incubated at 37°C for 30 min, followed by 2 min heat inactivation at 80°C.

The Cas9 ribonucleoprotein (RNP) was formed by combining 850ng of *in vitro* transcribed guide RNA pool, 1 µL of a 1:5 dilution of Alt-R *S.p*.Cas9 Nuclease V3 (1081058, IDT) or Alt-R *S.p*.HiFi Cas9 Nuclease V3 (1081060, IDT), and 1X rCutSmart buffer (B7204S, NEB) in a total of 30 µL. This reaction was incubated at room temperature for 20 min.

Next both the prepped gDNA and RNP are placed on ice before being combined. 1μL 10mMol dATP and 1.5μL Taq DNA Polymerase (M0273S, NEB) is added to the cut reaction and inverted and gently tapped to mix. This reaction is then incubated at 37°C for 30min for Cas9 cutting and brought to 75°C for a-tailing. CANVAS samples were additionally treated with 2ul of thermolabile Proteinase K (P8111S) for 15 min at 37°C, followed by heat inactivation at 55°C.

For adapter ligation, the cut reaction is transferred to a 1.5mL tube. We then added 5μL T4 DNA ligase (M0202M, NEB) and 5μL ONT LSK114 Ligation Adapter (LA; SQK-LSK114, ONT). This reaction is inverted to mix and incubated at room temperature for 20 min with rotation. Following ligation we add 1 volume of 1X TrisEDTA (TE) and invert to mix. Next 0.3X Ampure beads (SQK-LSK114, ONT) are added and incubated for 5 min with rotation followed by 5 min sitting at room temperature without rotation. The beads are then washed twice with 150μL Long Fragment Buffer (LFB; SQK-LSK114, ONT) followed by incubation with 20-50μL Elution Buffer (EB; SQK-LSK114, ONT) at 37°C for 30 min. Finally, we loaded the R10.4.1 MinION flow cell following the ONT protocol using 12μL of the library and sequenced for 72hrs.

### nCATs Data processing

ONT targeted sequencing data was basecalled and aligned using Dorado 0.6.0 using the super accuracy model with CG methylation calling.

## Results

To address the current need for comprehensive screening of disease-associated tandem repeat genotypes from nanopore data, we first developed a tandem repeat copy number caller to account for read level nanopore-specific error profiles, HMMSTR, as a companion bioinformatic tool for a targeted sequencing panel targeting 60 loci known or suspected of repeat expansion. We benchmark HMMSTR against four repeat plasmid constructs as well as two assemblies and compare its performance against current signal- and sequence-based methods compatible with targeted genotyping. Further, we demonstrate our sequencing strategy’s performance across three panels in two control cell lines and apply it to samples from nine individuals with either cerebellar ataxia, neuropathy, and vestibular areflexia syndrome (CANVAS) or amyotrophic lateral sclerosis/frontotemporal dementia (ALS/FTD). Using this strategy, we are able to genotype disease-associated expansions at previously uncharacterized loci in these individuals.

### A modified profile HMM for tandem repeat genotype determination from nanopore targeted sequencing reads

HMMSTR is optimized for targeted sequencing data and, as such, it assumes the majority of sequencing reads correspond to a given target. HMMSTR takes read data and performs local alignment of unique flanking sequences to assign reads to the most likely target. Assigned reads are then passed to the corresponding target and strand specific model. HMMSTR uses a modified profile HMM encoding the unique flanking sequence of a given target and a single copy of the expected repeat motif (Figure 1A, Materials and Methods). A modified Viterbi algorithm then labels the most likely positions of insertions and deletions before calculating the copy number for a given read. Once the repeat copy numbers are calculated across all reads and targets, allele copy numbers are called for each target using either a Gaussian mixture model or kernel density estimation (Materials and Methods).

### Benchmarking HMMSTR genotypes

To assess the accuracy of HMMSTR, we benchmark against three ground truth sets: four repeat expansion plasmid constructs, a PacBio HiFi assembly from the Human Genome Structural Variation consortium (HGSVC)(28), and the CHM13 reference genome from the telomere-to-telomere (T2T) project (40).

We ran HMMSTR on four plasmid constructs with variable repeat motif and copy number inserts ranging from 16 to 153 copies, including plasmids with ‘AAAAG’ (16,31, and 61 copies), ‘AAGGG’ (16, 31, and 61 copies), ‘GGGGC’ (21 and 41 copies) and ‘CGG’ motifs (20,39,77,115, and 153 copies). We show high concordance with all expected repeat copy numbers with a mean absolute difference of 1.23 copies and standard error of 0.7 per allele across all constructs (Figure. 1B, Supplementary Figure 1). We observe moderate heterogeneity in constructs, which is likely due to repeat instability in bacterial culture.

We next queried a total of 11,035 regions with repeat motif composition over 80% from GM12878 ONT WGS data (41) and PacBio circular consensus sequencing (CCS) WGS data (42). We ran both HMMSTR and Straglr (17) on all regions. We chose to benchmark against Straglr as it is a comparable sequence-based long-read genotyper that is currently utilized by Oxford Nanopore in their EPI2ME STR expansion workflow (https://epi2me.nanoporetech.com/). After filtering out homopolymers, regions with less than two read coverage, and regions with 200bp of masked sequence flanking the repeat of interest, the final number of regions genotyped by both HMMSTR and Straglr was 4,407 and 3,350 for PacBio and ONT respectively. We compare the correlation between these calls and the assembly in an allele specific manner (Methods). We demonstrate that HMMSTR (Figure 1C and D) outperforms Straglr (Figure 1E and F) in both our ONT and PacBio datasets (Supplementary Figure 2). The largest difference between our results in both datasets is the number of regions assigned to the haplotype 2. Of note, haplotype 1 includes the majority of homozygous calls and haplotype 2 includes the smaller alleles resulting from heterozygous calls. Straglr only calls 534 H2 regions compared to HMMSTR’s 1,240. Similar findings are observed in the PacBio results with Straglr assigning 316 alleles to the haplotype 2 set compared to 1,542 called by HMMSTR.

### Benchmarking heterozygous genotype calls

To validate HMMSTR heterozygous calls, we created a set of 19,256 heterozygous regions with at least a three copy difference between alleles and with non-repetitive flanking sequence (unmasked in the hg38 reference genome) from the PacBio HiFi assembly. Here, we find that HMMSTR has a higher correlation with the assembly compared to Straglr in heterozygous regions for both the ONT (Figures 2A and B 97% vs 64% PearsonR correlation for haplotype 1 set, Figure 2C and D 81% vs 45% PearsonR correlation for haplotype 2 set) and PacBio datasets (Supplementary Figures 3A and B 93% vs 66% PearsonR correlation for haplotype 1 set, Figures 2C and D 97% vs 78% PearsonR correlation for haplotype 2 set). As all the regions in this set are heterozygous in the assembly, we expect the number of regions assigned to both haplotype sets to be equal. To assess the concordance of our heterozygous calls with the assembly we calculate the number of regions called as heterozygous across each of our sets and the assembly. HMMSTR successfully calls 13,981/18,911 (74%) regions heterozygous in both datasets and the assembly with an increase in the number of heterozygous calls when the PacBio results are considered alone (94%)(Figure 2E). In contrast, Straglr calls 825/12,363 (6.7%) regions as heterozygous across all datasets (Figure 2F).

**Figure 2.**
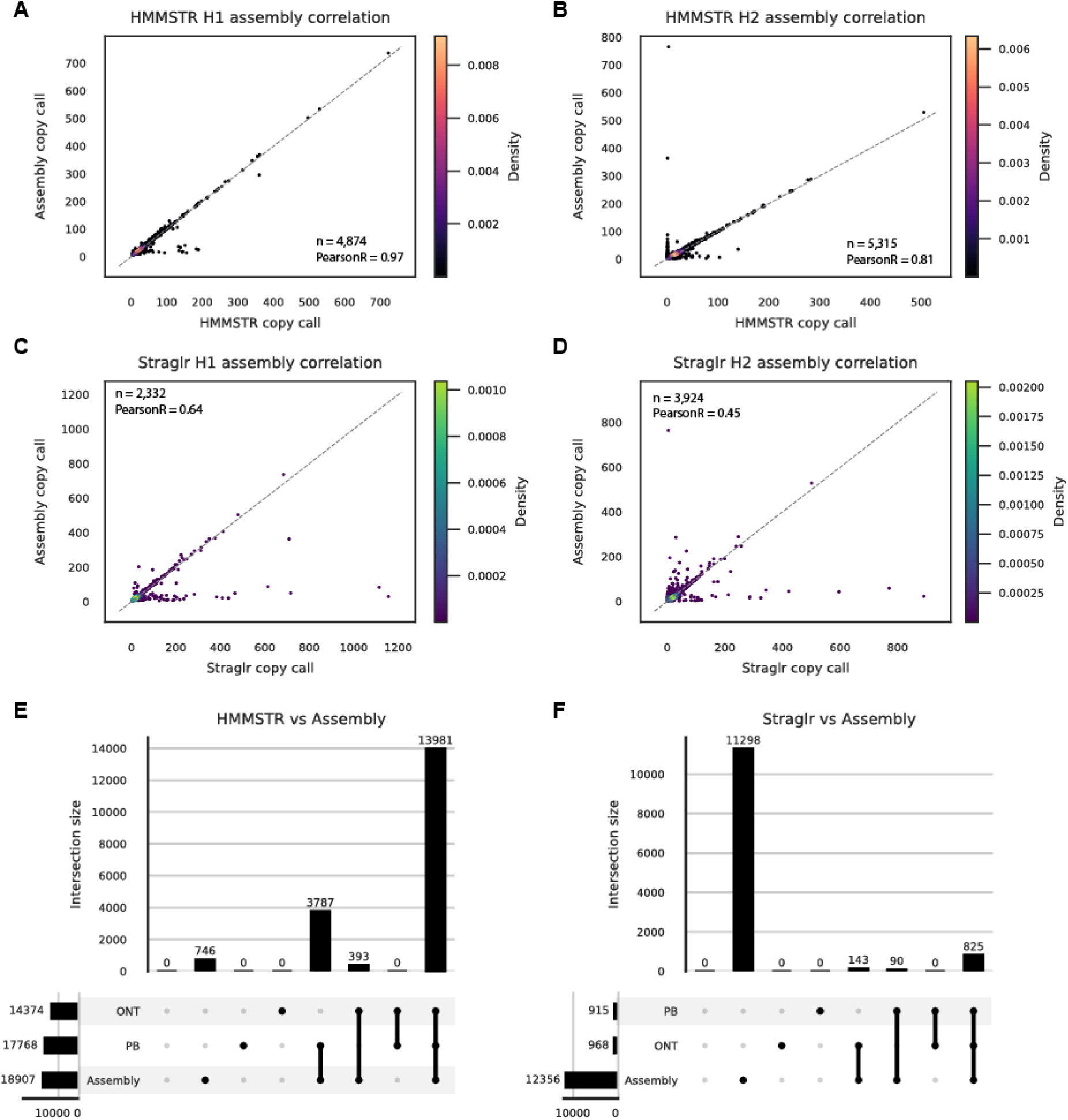
Heterozygous concordance with the assembly. (**A** and **B**). Correlation between HMMSTR copy number calls and HiFi assembly TRF estimates from heterozygous regions with at least a 3 motif copy number difference between alleles in the same ONT GM12878 dataset. **A.** larger allele correlation (n=4,874) and (**B**) smaller allele correlation (n=5,315). (**C** and **D**) Correlation between Straglr copy number calls and HiFi assembly TRF estimates for heterozygous regions in the same dataset. (**C**) larger allele correlation (n=2,332) and (**D**) smaller allele correlation (n=3,924). (**E** and **F**) Upset plots showing the number of regions called heterozygous across the ONT GM12878 dataset, PacBio GM12878 dataset, and the HiFi assembly. (**E**) HMMSTR call results and (n=18,907 loci). (**F**) Straglr call results (n=12,356 loci)

Chiu et al. state that Straglr resolves heterozygous tandem repeat loci when separated by over 100bp, thus we separated the regions by over or under 100bp difference. To note, when we subset assembly tandem repeats to only include heterozygous regions, we find that 98.9% of regions we identified from the simple repeats track differed by less than 100bp in the GM12878 assembly. We find that Straglr and HMMSTR call a comparable percentage of heterozygous regions across all datasets in the over 100bp group (82% Straglr vs 81% HMMSTR, n = 694 and n = 771 regions respectively) (Supplementary Figures 4C and D); however HMMSTR calls significantly more heterozygous genotypes than Straglr in regions with less than a 100bp difference (74% vs 2.2%, n= 18,136 and n= 11,662) (Supplementary Figures 4A and B).

### CHM13 benchmark

Next, we benchmarked HMMSTR and Straglr against a previously published short tandem repeat region set from the CHM13 reference genome (9). CHM13 is known to be effectively haploid (40), thus HMMSTR and Straglr were run with a maximum allele count of one. HMMSTR reports genotypes based on either the mode or median of a given read distribution, here we report results from both calls separately. We show HMMSTR calls CHM13 short tandem repeats with high accuracy (1.1 average copy difference from mode call, Figure 3A, 1.45 from median call, Supplementary Figure 5A) and Straglr calls the set with an average absolute difference of 4.97 (Supplementary Figure 5B). This is compared to the previously benchmarked callers DeepRepeat (9), RepeatHMM (43), and STRique (14) with 3.56, 4.47, and 11.2 average absolute differences respectively (9). Notably, both RepeatHMM and STRique share a similar profile HMM structure to HMMSTR, and DeepRepeat and STRique are both signal-based methods that were run on the same dataset. The authors note that they did not run either STRique or DeepRepeat on the full 126x dataset due to storage limitations; however, we show that we retain lower absolute copy difference in our downsampled set even at 5x average coverage (2.90 average absolute difference from mode call, 1.87 from median call, Figure 3B). Thus, not only does HMMSTR allow for the use of sequence data, it outperforms both current signal-based and profile HMM-based methods at low coverage.

**Figure 3.**
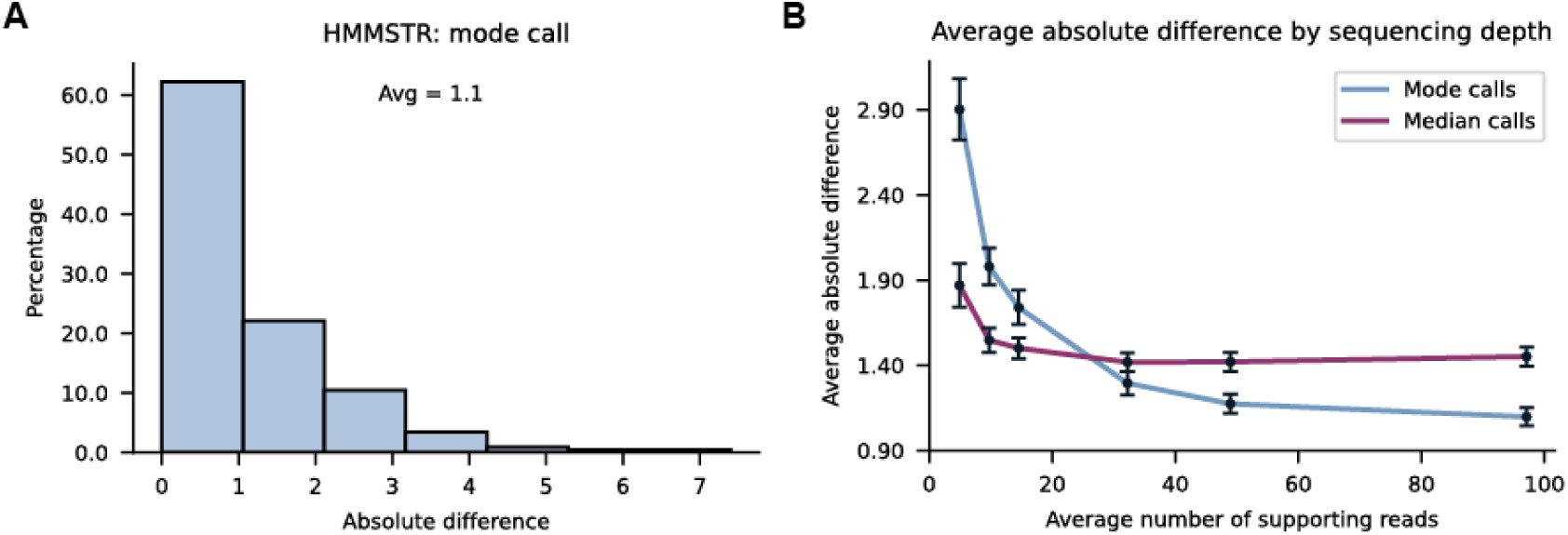
CHM13 HMMSTR Benchmark. (**A**) Average absolute difference between 439 long STR loci in CHM13 and HMMSTR mode calls from nanopore sequencing of CHM13. (**B**) Average absolute difference between CHM13 and HMMSTR median and mode calls from downsampled CHM13 nanopore dataset (5x-100x average coverage). Error bars show standard error per coverage tested.

### Multiplexed tandem repeat targeted sequencing

We next designed a set of flanking sgRNA guides targeting the majority of known disease-associated tandem repeat loci obtained through literature search (2, 29–33). These guides can be amplified and transcribed in a pooled approach that offers a simple and flexible multiplexed protocol. This set of guides was then used in the nCATSs approach to perform benchmark enrichments using a control sample with non-pathogenic repeat copy numbers at all loci (see Materials and Methods for details).

Using high molecular weight (HMW) genomic DNA (gDNA) from GM12878 lymphoblasts, we obtained an average coverage of 250x across 54 targets (Panel 1, Figure 4A). During guide optimization we designed an additional two panels (Panel 2 and Panel 3) where we added disease-associated targets and switched to IDT HiFi Cas9. This improved our on-target rates, where on-target is defined as the number of reads spanning the target tandem repeat loci divided by the total number of passing (Q>9) reads (Figure 4A, Supplementary Table 8).

**Figure 4.**
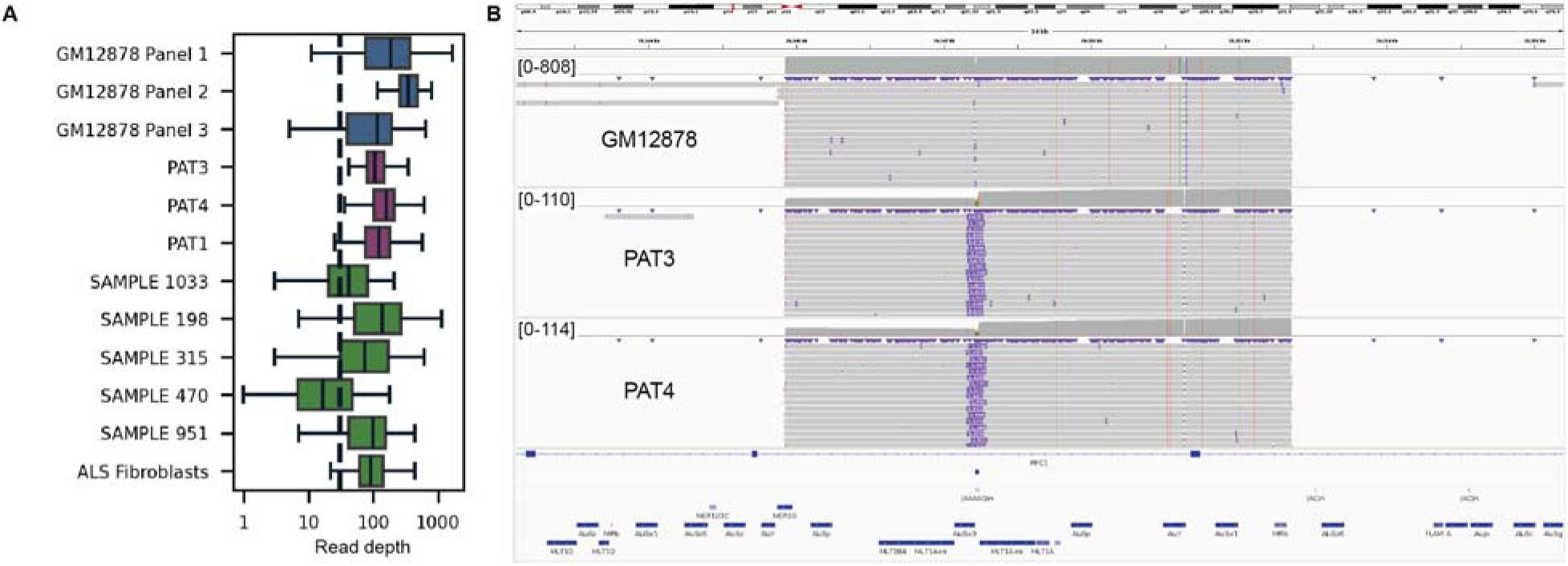
Targeted sequencing panel obtains high coverage over disease-associated STR. (**A**) Average read depth (log10 scale) at targets in 12 samples, including our control cell line GM12878, three CANVAS patient derived iPSC lines,, six post-mortem cerebellum samples from individuals with ALS/FTD, and one ALS patient-derived fibroblast line. (**B**) Integrative Genomics Viewer (IGV) image showing a 6.8kb region excised with flanking guides at *RFC1.* We obtained 808x read depth at this locus in the GM12878 control with no repeat expansion and 110x and 114x read depth in the CANVAS PAT3 and PAT4, both with large biallelic expansions.

### Characterization of CANVAS patient-derived iPSCs

After evaluating the targeted sequencing panel in a control cell line, we applied Panel 3 to three CANVAS patient-derived iPSC lines which carry large biallelic expansions in *RFC1* (PAT3 and PAT4). The autosomal recessive CANVAS expansion in intron two of *RFC1* is highly heterogeneous and multiple pathogenic and non-pathogenic alleles have been described (20, 44). While the reference, and most common allele, at this locus is made of up to 400 copies of the ‘AAAAG’ motif, pathogenic expansions include ‘AAGGG’, ‘ACAGG’, and ‘AGGGC’ motifs which can exceed 5kb in length (20, 44). Thus, long-read sequencing is particularly well suited to characterizing this locus.

Using the disease-associated panel to characterize these three lines, we obtained an average of 142x, 117x, and 168x read depth at our target repeat loci (Figure 4A). We observe biallelic ‘AAGGG’ expansions at the CANVAS locus in all three samples, with PAT3 having alleles with 821 and 947 copies, PAT4 harboring 1095 and 1174 copies, and PAT1 with 375 and 1228 copies (Figure 4B). In addition to spanning reads, we can also observe the large repeat expansions directly in soft clipped alignments and detect pathogenic length expansions in these fragmented reads (Supplementary Figure 10).

In the PAT1 sample, we observe not only a biallelic expansion at *RFC1*, but also a pathogenic length expansion at *FGF14* of 319 copies of the uninterrupted ‘AAG’ motif (Supplementary Figure 8). Interestingly, in PAT4, we also observe two intermediate sized *FGF14* expansions (146 and 211 copies) along with the biallelic *RFC1* expansions.

### Survey of ALS/FTD tissue samples for pathogenic repeat expansions

We next applied the 60 target panel (Panel 3) to six samples from individuals with neurodegenerative disease. A heterozygous “GGGGCC” repeat expansion in *C9orf72* is the most common genetic cause of ALS/FTD cases (45). Like the CANVAS expansion, the length of the pathogenic, expanded allele can exceed 5kb while the CG rich repeat motif can make characterization with amplification based methods more difficult (3, 46). Normal copies of the hexanucleotide motif range from 3-24 while pathogenic expansions can exceed one thousand copies (32, 33, 45). In addition, recent work has uncovered additional repeat expansions in NIPA1 and ATXN2 in individuals with ALS/FTD, suggesting potential pleiotropy in pathogenesis(45).

First, we used our method to characterize an ALS patient-derived cell line with a known *C9orf72* expansion. With our targeted sequencing panel we obtained high coverage at the locus and characterized the heterozygous expansion with HMMSTR (Figure. 5B). In this line, the pathogenic expanded allele contained 972 copies of the motif and the normal allele was nine copies (Figure 5A). In addition, in this sample we observed a heterozygous expansion in the *RFC1* intron which causes CANVAS. This expanded allele harbors 1487 copies of the pathogenic “AAGGG” motif and indicates this individual is a carrier for CANVAS (Figure 5B).

**Figure 5.**
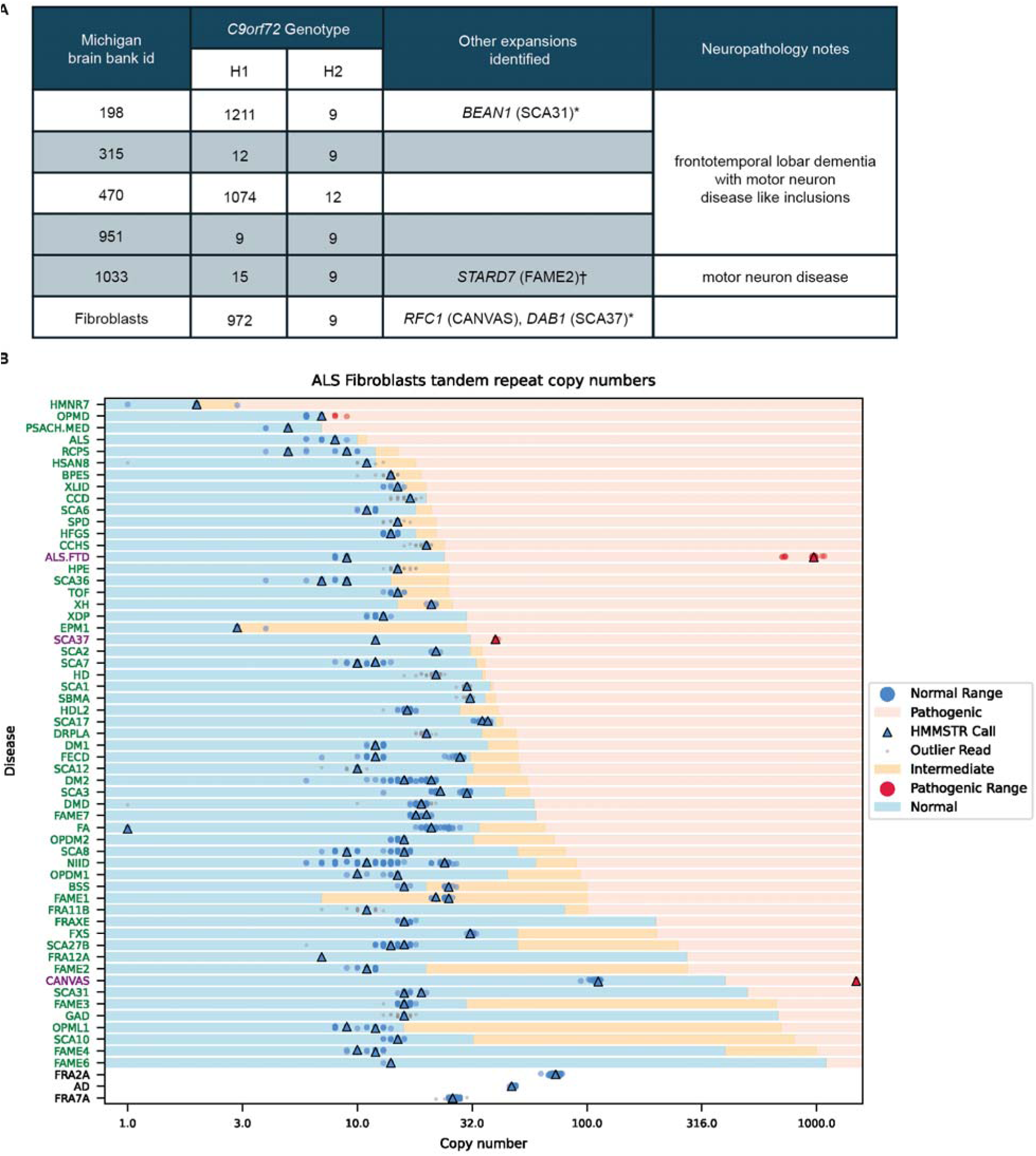
Application of targeted sequencing panel and genotyping with HMMSTR to ALS/FTD samples. (**A**) Sample information and *C9orf72* genotypes in five post-mortem cerebellum samples from individuals with ALS/FTD and one ALS patient-derived fibroblast line. Three samples carry one expanded *C9orf72* hexanucleotide repeat allele. In addition, we identified four expanded alleles. Patient 198 has a 536 copy number heterozygous, non-pathogenic repeat expansion in SCA31-associated *BEAN1*. Patient 1033 carries an expanded copy of the *STARD7* with 552 non-reference motifs (Supplementary Figure 10B) and the ALS/FTD fibroblast line carries one pathogenic CANVAS allele. (**B**) The swimlane plot shows the genotypes across 60 disease associated target loci for the ALS/FTD fibroblast line. All markers and ranges are displayed in log10 scale while the x-axis reflects the absolute copy number. Dots indicate repeat counts for each read and triangles show HMMSTR median calls, where blue markings correspond to normal allele sizes and red indicate pathogenic length reads and calls. Gray dots indicate outlier copy number calls. Stars show fragmented reads that didn’t span the repeat (soft clipped in alignment). For each disease associated locus, the log copy number x-axis has been shaded to show the ranges of normal, intermediate, and pathogenic repeat copy numbers. For rows without shading, the ranges of normal and pathogenic lengths have not been described. Disease abbreviations (y-axis) shown in green indicate HMMSTR call was in the normal range for both alleles, purple indicates one allele was in the pathogenic range, and black indicates missing data or no ranges available(32, 33). *non-pathogenic motif. †motif of unknown significance.

Next we genotyped six post-mortem cerebellum samples where we identified two heterozygous *C9orf72* expansions. Overall, due to HMW gDNA quality, the tissue samples yielded lower coverage than experiments using cell lines, however we were able to successfully genotype most target loci. To investigate the possibility of other expansion loci contributing to disease, we examined the genotypes of these samples at the rest of the disease-associated tandem repeat loci. We identified two additional interesting repeat expansions in these individuals (Supplemental Figure 10). In the sample from individual 198, in addition to an expanded *C9orf72* allele, we observed a heterozygous expansion in the spinocerebellar ataxia 31 (SCA31) associated repeat in *BEAN1*. The pathogenic expansion at this locus is a non-reference, nested repeat expansion consisting of ‘AAAAT’, ‘AAGGT,’ and ‘AAGAT’(Supplementary Figure 10A). In this sample we obtained an inconclusive genotype with approximately 528 copies of an “AAAAT”, “AAGAT”, or “AATGG” motif that was inconsistent in expanded reads.

We also identified a potentially pathogenic expansion in individual 1033 at the familial adult myoclonic epilepsy 2 (FAME2) associated locus in *STARD7*. Like SCA31 there are pathogenic and non-pathogenic motifs. FAME loci are heterogeneous and several additional motifs have been identified at this locus across populations, but the pathogenicity of some of these motifs is still unclear (47). In this sample, we observed a 552 copy heterozygous expansion of the uncharacterized motif, ‘AATAC’.

In addition to the expansions we identified in these samples, we also observed a number of intermediate expansions as well as rare and non-reference alleles that would have been missed using short-read sequencing. Examples include large, reference alleles at FAME loci, and a previously described rare, complex allele at *RFC1* (Supplementary Figure 6A)(48).

## Discussion

Accurate genotyping is essential for understanding and diagnosing disease-associated tandem repeat expansions. Here we establish a computational method designed for targeted sequencing, HMMSTR, that outperforms both signal-based and sequence-based tandem repeat copy number callers and provide a flexible and robust targeted sequencing panel for disease-associated tandem repeats. To demonstrate HMMSTR’s performance, we benchmark against repeat constructs and assembly data and show high concordance across all sets for both ONT and PacBio HiFi datasets.

Paired with targeted sequencing, HMMSTR’s sequence-based approach is efficient and requires fewer resources for storage than WGS or signal-based genotyping. It further outperforms these methods in accurately discriminating between similarity sized, heterozygous repeats. Though the accuracy of ONT sequencing has improved significantly, sequencing errors in low complexity regions still impede direct genotyping, particularly at low coverage. HMMSTR models repeat errors to address this and is able to call copy numbers at as low as 5x coverage.

HMMSTR is optimized for targeted sequencing and assumes there are minimal off-target reads. However, HMMSTR’s use of local alignment introduces potential for mis-assigning reads when there are a large number of off-target reads or targets with similar flanking sequence. This limitation is particularly relevant when attempting to run HMMSTR on WGS data. To account for this in our benchmarking, we perform a global alignment and subset the data to only include our target regions to simulate a targeted sequencing run. As some tandem repeats are located within low-complexity regions, HMMSTR allows for longer flanking sequence input for both alignment and the model encoding to help with target specificity at the cost of runtime.

While one advantage of HMMSTR’s targeted approach is its specificity and ability to account for errors based on prior knowledge of target loci, challenges may persist when considering the variability of some disease-associated tandem repeats. Previous studies have reported over 98% of tandem repeats have low sequence polymorphism (20), but multiple disease-associated tandem repeats carry non-reference motifs or complex motif composition. Models constructed on reference motifs generally obtain the same copy number as models constructed with pathogenic motifs due to the similarity in both length and base composition, as well as specificity of flanking sequences. However, we do find that in some expansion cases with non-reference motifs, HMMSTR may underestimate the copy number using a reference motif model. Motif composition can be recovered in downstream analysis from the per-read repeat coordinates returned by HMMSTR given adequate coverage (Supplementary Figure 6). To account for the shift in copy number due to non-reference motif composition in the expansion cases described in our analysis, HMMSTR was run with reference and pathogenic motifs when applicable. Integration of alternate motifs into target models would potentially increase the efficiency and ease-of-use of genotyping these variable regions and future iterations of HMMSTR may incorporate these optimizations. (20). Thus, HMMSTR is broadly applicable when using expected repeat motifs for copy number determination and copy numbers may be refined by inclusion of known pathogenic motifs.

After benchmarking HMMSTR we were able to successfully apply our targeted sequencing panel and genotyping to a variety of samples, including two patient groups. Using patient-derived lines and post-mortem cerebellum samples, we are able to genotype large, biallelic repeat expansions at the CANVAS locus in *RFC1* and large, CG rich expansions in *C9orf72*.

The rate of co-occurrent repeat expansion in the samples we tested highlights the utility of this targeted panel approach. Here, we are able to investigate repeat expansions at other disease associated loci in addition to characterizing CANVAS and ALS/FTD expansions in the corresponding samples.

In two of the three CANVAS samples, we identified potentially pathogenic expansions in *FGF14*, another common cause of late onset cerebellar ataxia with overlapping disease presentation (49, 50). Few cases of SCA27B have been reported in CANVAS carriers (51), however to our knowledge this is the first reported biallelic *RFC1* and *FGF14* expansion to date. In PAT4, we observe two intermediate sized *FGF14* expansions with biallelic *RFC1* expansions. Recent efforts to better characterize the *FGF14* expansion associated with SCA27B have suggested that biallelic, intermediate, expansions where alleles do not exceed the full penetrance threshold of 300 copies may exhibit an additive effect to determine penetrance (49, 52). While investigation into the effects of these *RFC1* and *FGF14* co-expansions exceeds the scope of this current work, future application of our panel may elucidate if these co-occurences have an effect on disease etiology or presentation in affected individuals as well as aid in differential diagnosis of these two disorders.

Multiple pathogenic and intermediate tandem repeat expansions have been identified in individuals with ALS/FTD with and without the *C9orf72* expansion (45, 53). The prevalence and significance of these co-occurrent and additional expansions is yet to be determined, highlighting the importance of screening multiple loci (45). In the ALS/FTD samples we analyzed, we identified several expanded and non-reference alleles at the panel targets including in *STARD7, BEAN1, RFC1,* and *DAB1*. Recently whole genome, long-read sequencing projects have uncovered significant heterogeneity in both motif composition and size at these pentanucleotide repeat loci, as well as other disease-associated tandem repeats, across populations (20, 54). Using this targeted panel we are able to efficiently and accurately genotype these loci as well as rare and complex alleles with HMMSTR.This enables an increased power to investigate and characterize a wide range of variation at disease-associated tandem repeats in both cases and controls.

Despite our ability to target our repeats of interest with high coverage, some genotyping challenges remain. Though we observe very high coverage at guide cut sites with targeted sequencing, we obtain fewer reads that span very large expansions. This is consistent with other work and may be due to fragmentation and secondary structure, particularly in samples with pathogenic motifs (15).

Overall, this pooled guide strategy for genotyping disease-associated tandem repeat expansions offers a simple, flexible, and accurate screening methods for interrogating a wide array of samples and targets. In addition, this method is highly scalable, as amplification and transcription of the pool can yield sufficient sgRNA for hundreds of samples. Since the design of Panel 3, recent work has identified and described additional disease-associated repeats and we aim to iteratively optimize our panel and add targets as necessary for comprehensive screening (33, 55, 56). Moreover, as FDA approved treatments emerge for repeat expansion disorders and they become the subject of gene-targeting clinical trials, critical additional information related to polymorphic repeat structures and alterations in surrounding sequence will prove critical for accurate management of therapeutic options.

In conclusion, we demonstrate the utility of our combined targeted bioinformatic and sequencing strategy. By obtaining high coverage and accurate genotypes across disease-associated loci simultaneously, we are able to not only genotype normal-length repeats and confirm pathogenic expansions, but also identify expansions at unexpected loci and co-expansions in samples from individuals with neurodegenerative conditions. This strategy holds promise for more economic and comprehensive diagnostics as well as further study of the diversity at previously elusive tandem repeat loci.

## Acknowledgements

We thank members of the Todd, Mills, and Boyle labs for helpful discussions and suggestions related to his manuscript. We would like to acknowledge Mr. Matthew D. Perkins for his help with postmortem tissue from the University of Michigan Brain Bank. In addition, we would like to thank the generous patients who contributed their tissues.

## Author Contributions

A.P.B., P.K.T., K.V., and C.M. conceived the project. C.J.M. and J.S. established and cultured cell lines and isolated gDNA. P.K.T and C.J.M obtained patient tissue samples for culture and screening. C.M. developed the guide panels and performed Cas9 targeted enrichment and nanopore sequencing. K.V. developed and benchmarked HMMSTR as well as performed computational analysis. All authors guided the data analysis strategy. A.P.B., K.V., and C.M. wrote the manuscript. All authors edited the manuscript. All authors read and approved the final manuscript.

## Data Availability

Data from GM12878 and ALS/FTD Cas9 Enrichments are available at SRA bioproject PRJNA1079777. Additional sample data is available upon reasonable request.

## Code Availability

HMMSTR is available at https://github.com/Boyle-Lab/HMMSTR.

## References

1. English, A.C., Dolzhenko, E., Ziaei Jam, H., McKenzie, S.K., Olson, N.D., De Coster, W., Park, J., Gu, B., Wagner, J., Eberle, M.A., et al. (2024) Analysis and benchmarking of small and large genomic variants across tandem repeats. Nat. Biotechnol.

2. Malik, I., Kelley, C.P., Wang, E.T. and Todd, P.K. (2021) Molecular mechanisms underlying nucleotide repeat expansion disorders. Nat. Rev. Mol. Cell Biol., 22, 589–607.

3. Ciotti, P., Di Maria, E., Bellone, E., Ajmar, F. and Mandich, P. (2004) Triplet Repeat Primed PCR (TP PCR) in Molecular Diagnostic Testing for Friedreich Ataxia. J. Mol. Diagn., 6, 285.

4. Ibañez, K., Polke, J., Tanner Hagelstrom, R., Dolzhenko, E., Pasko, D., Thomas, E.R.A., Daugherty, L.C., Kasperaviciute, D., Smith, K.R., McDonagh, E.M., et al. (2022) Whole genome sequencing for the diagnosis of neurological repeat expansion disorders in the UK: a retrospective diagnostic accuracy and prospective clinical validation study. Lancet Neurol., 21, 234–245.

5. Dolzhenko, E., Deshpande, V., Schlesinger, F., Krusche, P., Petrovski, R., Chen, S., Emig-Agius, D., Gross, A., Narzisi, G., Bowman, B., et al. (2019) ExpansionHunter: a sequence-graph-based tool to analyze variation in short tandem repeat regions. Bioinformatics, 35, 4754–4756.

6. Dolzhenko, E., Bennett, M.F., Richmond, P.A., Trost, B., Chen, S., van Vugt, J.J.F.A., Nguyen, C., Narzisi, G., Gainullin, V.G., Gross, A.M., et al. (2020) ExpansionHunter Denovo: a computational method for locating known and novel repeat expansions in short-read sequencing data. Genome Biol., 21, 1–14.

7. Dashnow, H., Pedersen, B.S., Hiatt, L., Brown, J., Beecroft, S.J., Ravenscroft, G., LaCroix, A.J., Lamont, P., Roxburgh, R.H., Rodrigues, M.J., et al. (2022) STRling: a k-mer counting approach that detects short tandem repeat expansions at known and novel loci. Genome Biol., 23, 1–20.

8. Mousavi, N., Shleizer-Burko, S., Yanicky, R. and Gymrek, M. (2019) Profiling the genome-wide landscape of tandem repeat expansions. Nucleic Acids Res., 47, e90.

9. Fang, L., Liu, Q., Monteys, A.M., Gonzalez-Alegre, P., Davidson, B.L. and Wang, K. (2022) DeepRepeat: direct quantification of short tandem repeats on signal data from nanopore sequencing. Genome Biol., 23, 108.

10. Delahaye, C. and Nicolas, J. (2021) Sequencing DNA with nanopores: Troubles and biases. PLoS One, 16, e0257521.

11. Oehler, J.B., Wright, H., Stark, Z., Mallett, A.J. and Schmitz, U. (2023) The application of long-read sequencing in clinical settings. Hum. Genomics, 17, 1–13.

12. Stevanovski, I., Chintalaphani, S.R., Gamaarachchi, H., Ferguson, J.M., Pineda, S.S., Scriba, C.K., Tchan, M., Fung, V., Ng, K., Cortese, A., et al. (2022) Comprehensive genetic diagnosis of tandem repeat expansion disorders with programmable targeted nanopore sequencing. Sci Adv, 8, eabm5386.

13. Sitarčík, J., Vinař, T., Brejová, B., Krampl, W., Budiš, J., Radvánszky, J. and Lucká, M. (2023) WarpSTR: determining tandem repeat lengths using raw nanopore signals. Bioinformatics, 39.

14. Giesselmann, P., Brändl, B., Raimondeau, E., Bowen, R., Rohrandt, C., Tandon, R., Kretzmer, H., Assum, G., Galonska, C., Siebert, R., et al. (2019) Analysis of short tandem repeat expansions and their methylation state with nanopore sequencing. Nat. Biotechnol., 37, 1478–1481.

15. Erdmann, H., Schöberl, F., Giurgiu, M., Leal Silva, R.M., Scholz, V., Scharf, F., Wendlandt, M., Kleinle, S., Deschauer, M., Nübling, G., et al. (2022) Parallel in-depth analysis of repeat expansions in ataxia patients by long-read sequencing. Brain, 146, 1831–1843.

16. Gilpatrick, T., Lee, I., Graham, J.E., Raimondeau, E., Bowen, R., Heron, A., Downs, B., Sukumar, S., Sedlazeck, F.J. and Timp, W. (2020) Targeted nanopore sequencing with Cas9-guided adapter ligation. Nat. Biotechnol., 38, 433–438.

17. Chiu, R., Rajan-Babu, I.-S., Friedman, J.M. and Birol, I. (2021) Straglr: discovering and genotyping tandem repeat expansions using whole genome long-read sequences. Genome Biol., 22, 224.

18. Mitsuhashi, S., Frith, M.C., Mizuguchi, T., Miyatake, S., Toyota, T., Adachi, H., Oma, Y., Kino, Y., Mitsuhashi, H. and Matsumoto, N. (2019) Tandem-genotypes: robust detection of tandem repeat expansions from long DNA reads. Genome Biol., 20, 58.

19. Bolognini, D., Magi, A., Benes, V., Korbel, J.O. and Rausch, T. (2020) TRiCoLOR: tandem repeat profiling using whole-genome long-read sequencing data. Gigascience, 9.

20. Dolzhenko, E., English, A., Dashnow, H., De Sena Brandine, G., Mokveld, T., Rowell, W.J., Karniski, C., Kronenberg, Z., Danzi, M.C., Cheung, W.A., et al. (2024) Characterization and visualization of tandem repeats at genome scale. Nat. Biotechnol.

21. Brais, B., Pellerin, D. and Danzi, M.C. (2023) Deep Intronic FGF14 GAA Repeat Expansion in Late-Onset Cerebellar Ataxia. Reply. N. Engl. J. Med., 388, e70.

22. Mumm, C., Drexel, M.L., McDonald, T.L., Diehl, A.G., Switzenberg, J.A. and Boyle, A.P. (2023) Multiplexed long-read plasmid validation and analysis using OnRamp. Genome Res., 33, 741–749.

23. Eddy, S.R. (1998) Profile hidden Markov models. Bioinformatics, 14, 755–763.

24. Baum, L.E., Petrie, T., Soules, G. and Weiss, N. (1970) A maximization technique occurring in the statistical analysis of probabilistic functions of Markov chains. Ann. Math. Stat., 41, 164–171.

25. Forney, G.D. (1973) The viterbi algorithm. Proc. IEEE Inst. Electr. Electron. Eng., 61, 268– 278.

26. Li, H. (2018) Minimap2: pairwise alignment for nucleotide sequences. Bioinformatics, 34, 3094–3100.

27. Benson, G. (1999) Tandem repeats finder: a program to analyze DNA sequences. Nucleic Acids Res., 27, 573–580.

28. Porubsky, D., Ebert, P., Audano, P.A., Vollger, M.R., Harvey, W.T., Marijon, P., Ebler, J., Munson, K.M., Sorensen, M., Sulovari, A., et al. (2021) Fully phased human genome assembly without parental data using single-cell strand sequencing and long reads. Nat. Biotechnol., 39, 302–308.

29. Repeat expansion diseases (2018) In Handbook of Clinical Neurology. Elsevier, Vol. 147, pp. 105–123.

30. 30 years of repeat expansion disorders: What have we learned and what are the remaining challenges? (2021) Am. J. Hum. Genet., 108, 764–785.

31. Chintalaphani, S.R., Pineda, S.S., Deveson, I.W. and Kumar, K.R. (2021) An update on the neurological short tandem repeat expansion disorders and the emergence of long-read sequencing diagnostics. Acta Neuropathologica Communications, 9, 1–20.

32. Halman, A., Dolzhenko, E. and Oshlack, A. (2022) STRipy: A graphical application for enhanced genotyping of pathogenic short tandem repeats in sequencing data. Hum. Mutat., 43, 859–868.

33. Chaisson, M.J.P., Sulovari, A., Valdmanis, P.N., Miller, D.E. and Eichler, E.E. (2023) Advances in the discovery and analyses of human tandem repeats. Emerg Top Life Sci, 7, 361–381.

34. Masutani, B., Kawahara, R. and Morishita, S. (2023) Decomposing mosaic tandem repeats accurately from long reads. Bioinformatics, 39, btad185.

35. Maltby, C.J., Krans, A., Grudzien, S.J., Palacios, Y., Muiños, J., Suárez, A., Asher, M., Khurana, V., Barmada, S.J., Dijkstra, A.A., et al. (2023) AAGGG repeat expansions trigger RFC1-independent synaptic dysregulation in human CANVAS Neurons. bioRxiv, 10.1101/2023.12.13.571345.

36. Labun, K., Montague, T.G., Krause, M., Torres Cleuren, Y.N., Tjeldnes, H. and Valen, E. (2019) CHOPCHOP v3: expanding the CRISPR web toolbox beyond genome editing. Nucleic Acids Res., 47, W171–W174.

37. Anthon, C., Corsi, G.I. and Gorodkin, J. (2022) CRISPRon/off: CRISPR/Cas9 on- and off-target gRNA design. Bioinformatics, 38, 5437–5439.

38. Gilpatrick, T., Wang, J.Z., Weiss, D., Norris, A.L., Eshleman, J. and Timp, W. (2023) IVT generation of guideRNAs for Cas9-enrichment Nanopore Sequencing. bioRxiv, 10.1101/2023.02.07.527484.

39. McDonald, T.L., Zhou, W., Castro, C.P., Mumm, C., Switzenberg, J.A., Mills, R.E. and Boyle, A.P. (2021) Cas9 targeted enrichment of mobile elements using nanopore sequencing. Nat. Commun., 12, 1–13.

40. Nurk, S., Koren, S., Rhie, A., Rautiainen, M., Bzikadze, A.V., Mikheenko, A., Vollger, M.R., Altemose, N., Uralsky, L., Gershman, A., et al. (2022) The complete sequence of a human genome. Science, 376, 44–53.

41. Jain, M., Koren, S., Miga, K.H., Quick, J., Rand, A.C., Sasani, T.A., Tyson, J.R., Beggs, A.D., Dilthey, A.T., Fiddes, I.T., et al. (2018) Nanopore sequencing and assembly of a human genome with ultra-long reads. Nat. Biotechnol., 36, 338–345.

42. Vollger, M.R., Dishuck, P.C., Harvey, W.T., DeWitt, W.S., Guitart, X., Goldberg, M.E., Rozanski, A.N., Lucas, J., Asri, M., Munson, K.M., et al. (2023) Increased mutation and gene conversion within human segmental duplications. Nature, 617, 325–334.

43. Liu, Q., Zhang, P., Wang, D., Gu, W. and Wang, K. (2017) Interrogating the ‘unsequenceable’ genomic trinucleotide repeat disorders by long-read sequencing. Genome Med., 9, 1–16.

44. Dominik, N., Magri, S., Currò, R., Abati, E., Facchini, S., Corbetta, M., Macpherson, H., Bella, D., Sarto, E., Stevanovski, I., et al. (2023) Normal and pathogenic variation of RFC1 repeat expansions: implications for clinical diagnosis. Brain, 146, 5060–5069.

45. Henden, L., Fearnley, L.G., Grima, N., McCann, E.P., Dobson-Stone, C., Fitzpatrick, L., Friend, K., Hobson, L., Chan Moi Fat, S., Rowe, D.B., et al. (2023) Short tandem repeat expansions in sporadic amyotrophic lateral sclerosis and frontotemporal dementia. Sci Adv, 9, eade2044.

46. Fazal, S., Danzi, M.C., Cintra, V.P., Bis-Brewer, D.M., Dolzhenko, E., Eberle, M.A. and Zuchner, S. (2020) Large scale in silico characterization of repeat expansion variation in human genomes. Scientific Data, 7, 1–14.

47. Corbett, M.A., Kroes, T., Veneziano, L., Bennett, M.F., Florian, R., Schneider, A.L., Coppola, A., Licchetta, L., Franceschetti, S., Suppa, A., et al. (2019) Intronic ATTTC repeat expansions in STARD7 in familial adult myoclonic epilepsy linked to chromosome 2. Nat. Commun., 10, 1–10.

48. Dolzhenko, E., English, A., Dashnow, H., De Sena Brandine, G., Mokveld, T., Rowell, W.J., Karniski, C., Kronenberg, Z., Danzi, M.C., Cheung, W., et al. (2023) Resolving the unsolved: Comprehensive assessment of tandem repeats at scale. bioRxiv, 10.1101/2023.05.12.540470.

49. Ouyang, R., Wan, L., Pellerin, D., Long, Z., Hu, J., Jiang, Q., Wang, C., Peng, L., Peng, H., He, L., et al. (2024) The genetic landscape and phenotypic spectrum of GAA-FGF14 ataxia in China: a large cohort study. eBioMedicine, 102.

50. Pellerin, D., Wilke, C., Traschütz, A., Nagy, S., Currò, R., Dicaire, M.-J., Garcia-Moreno, H., Anheim, M., Wirth, T., Faber, J., et al. (2024) Intronic GAA repeat expansions are a common cause of ataxia syndromes with neuropathy and bilateral vestibulopathy. J. Neurol. Neurosurg. Psychiatry, 95, 175–179.

51. Awad, P.S., Lohmann, K., Hirmas, Y., Hinrichs, F., Thomsen, M., Kauffman, M., Lüth, T., Trinh, J., Westenberger, A., Chaná-Cuevas, P., et al. (2023) Shaking Up Ataxia: FGF14 and RFC1 Repeat Expansions in Affected and Unaffected Members of a Chilean Family. Mov. Disord., 38, 1107–1109.

52. Depienne, C., Mohren, L., Erdlenbruch, F., Leitão, E., Kilpert, F., Sebastian Hönes, G., Thieme, A., Sturm, M., Park, J., Schlüter, A., et al. (2024) Unveiling pathogenic and non-pathogenic FGF14 repeat expansions: identification, sequence, and secondary structure. 10.21203/rs.3.rs-3940197/v1.

53. Nagy, Z.F., Pál, M., Engelhardt, J.I., Molnár, M.J., Klivényi, P. and Széll, M. (2024) Beyond C9orf72: repeat expansions and copy number variations as risk factors of amyotrophic lateral sclerosis across various populations. BMC Med. Genomics, 17, 1–8.

54. Gustafson, J.A., Gibson, S.B., Damaraju, N., Zalusky, M.P.G., Hoekzema, K., Twesigomwe, D., Yang, L., Snead, A.A., Richmond, P.A., De Coster, W., et al. (2024) Nanopore sequencing of 1000 Genomes Project samples to build a comprehensive catalog of human genetic variation. medRxiv, 10.1101/2024.03.05.24303792.

55. Exonic trinucleotide repeat expansions in ZFHX3 cause spinocerebellar ataxia type 4: A poly-glycine disease (2024) Am. J. Hum. Genet., 111, 82–95.

56. English, A., Dolzhenko, E., Jam, H.Z., Mckenzie, S., Olson, N.D., De Coster, W., Park, J., Gu, B., Wagner, J., Eberle, M.A., et al. (2023) Benchmarking of small and large variants across tandem repeats. bioRxiv, 10.1101/2023.10.29.564632.

