## Supplementary Data for "Enhanced Detection and Genotyping of Disease-Associated Tandem Repeats Using HMMSTR and Targeted Long-Read Sequencing"

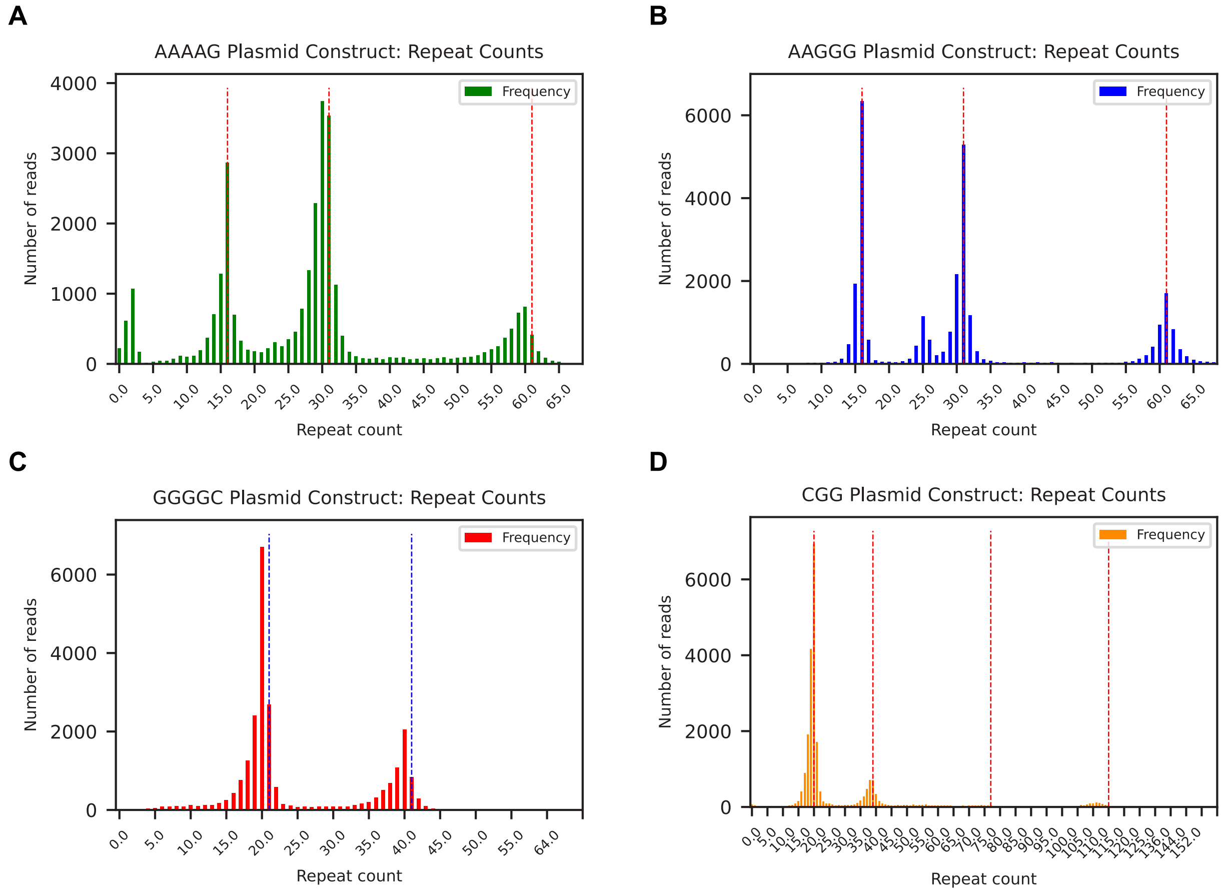


**Supplemental Figure 1: HMMSTR results from repeat containing plasmid constructs.**

(**A**) Results from AAAAG plasmid benchmarking construct with target repeat copy numbers 16, 31, and 61, HMMSTR calls 16, 30 and 60. (**B**) Results from AAGGG plasmid benchmarking construct with target repeat copy numbers 16, 31, and 61, HMMSTR calls 16, 31, and 61. (**C**) Results from GGGGC plasmid benchmarking construct with target repeat lengths 21 and 41, HMMSTR calls 20 and 40. (**D**) Results from CGG plasmid benchmarking construct with target repeat copy numbers 20, 39, 77, 115, and 153, HMMSTR calls 20, 39, 76, 111, 144.

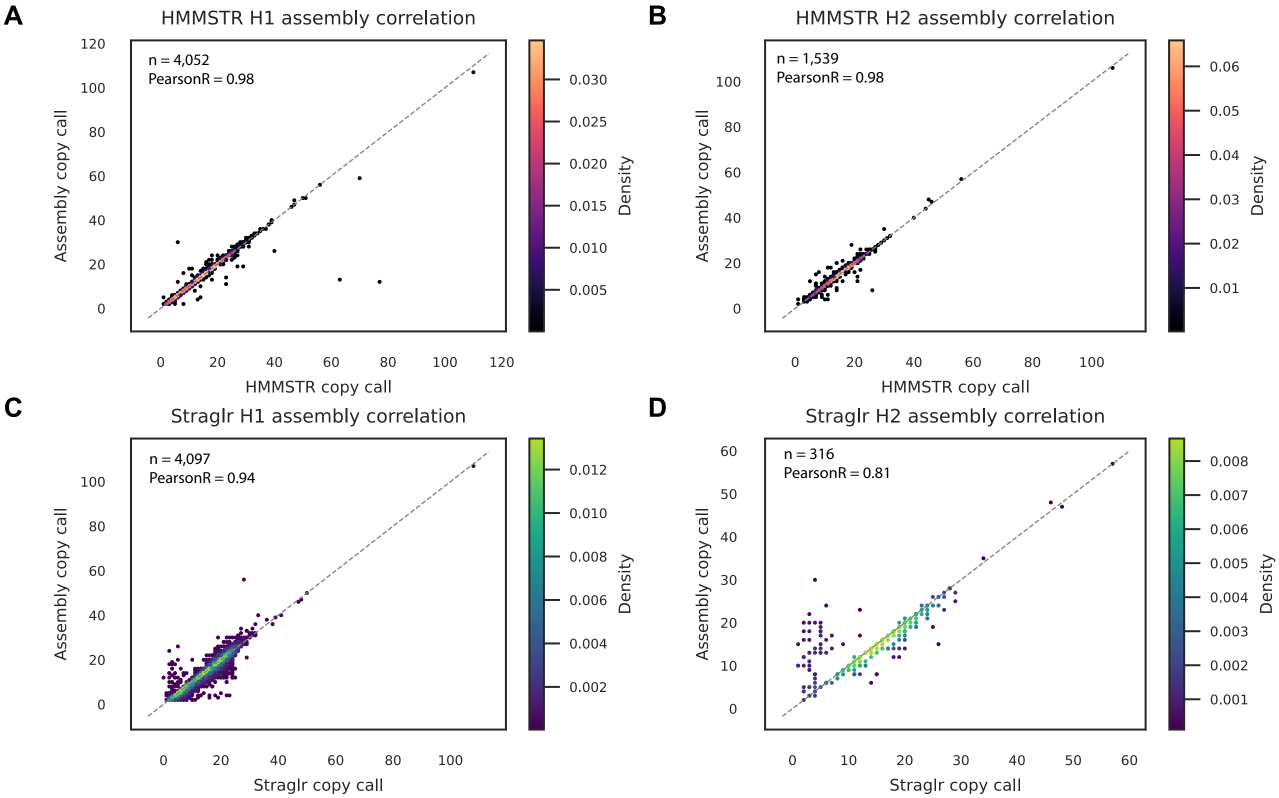


**Supplemental Figure 2: HMMSTR and Straglr concordance with ground truth sets: PacBio CCS dataset vs Assembly.**

(**A** and **B**) Correlation between HMMSTR copy number calls from GM12878 PacBio CCS dataset and HiFi assembly copynumbers estimated by TRF on GM12878. (**A)** Correlation between homozygous loci and largest allele calls (n=4,052 loci) and (**B)** correlation between smallest allele calls (n=1,539). (**C** and **D**) Correlation between Straglr copy number calls from the same PacBio CCS dataset, assembly, and regions copy number estimates. (**C**) Correlation between homozygous loci and largest allele calls (n=4,097 loci) and (**D**) correlation between smallest allele calls (n=316).

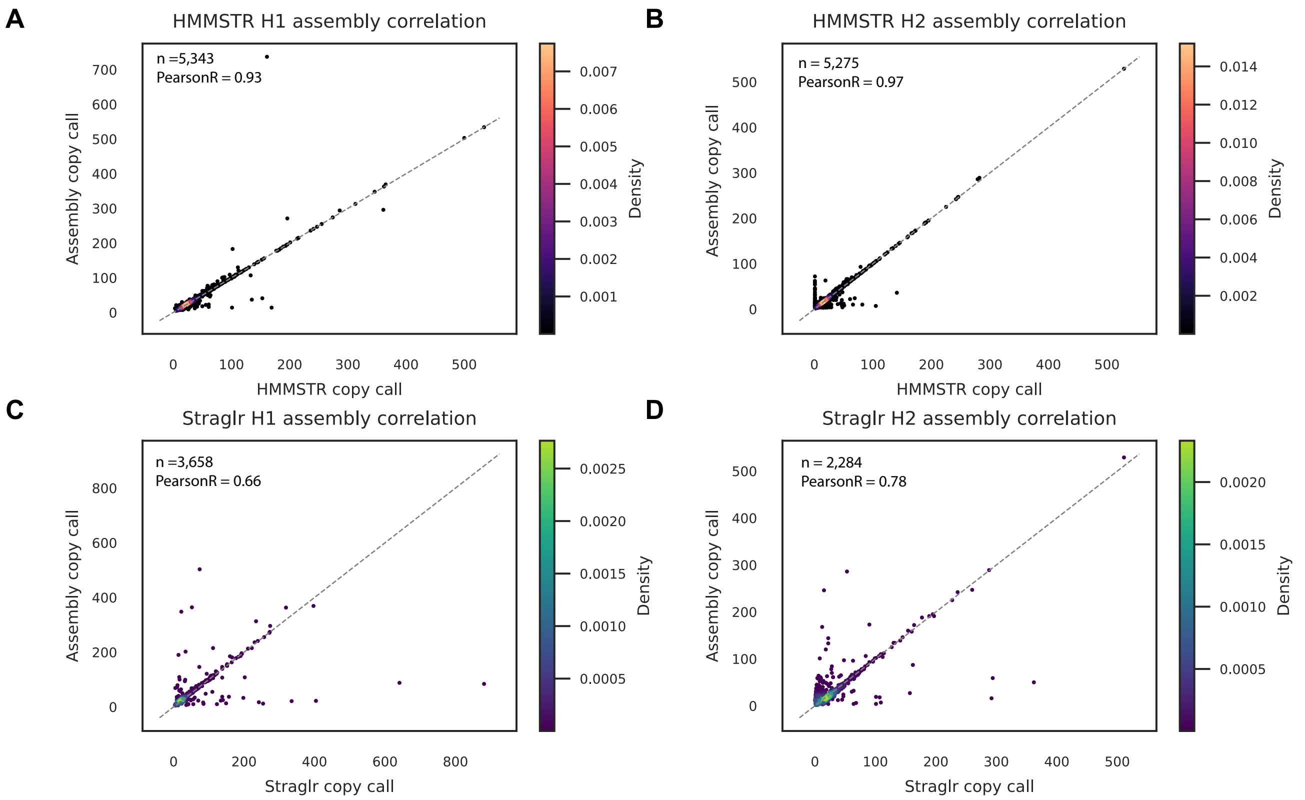


**Supplemental Figure 3. Heterozygous concordance with the assembly: HMMSTR vs Straglr.**

(**A** and **B**) Correlation between HMMSTR copy number calls and HiFi assembly TRF estimates from heterozygous regions with at least a 3 motif copy number difference between alleles in the PacBio CCS GM12878 dataset. (**A**) larger allele correlation (n=5,343) and (**B**) smaller allele correlation (n=5,275) (**C** and **D**) Correlation between Straglr copy number calls and HiFi assembly TRF estimates for the same heterozygous regions in the same dataset. (**C**) larger allele correlation (n=3,658) and (**D**) smaller allele correlation (n=2,284).

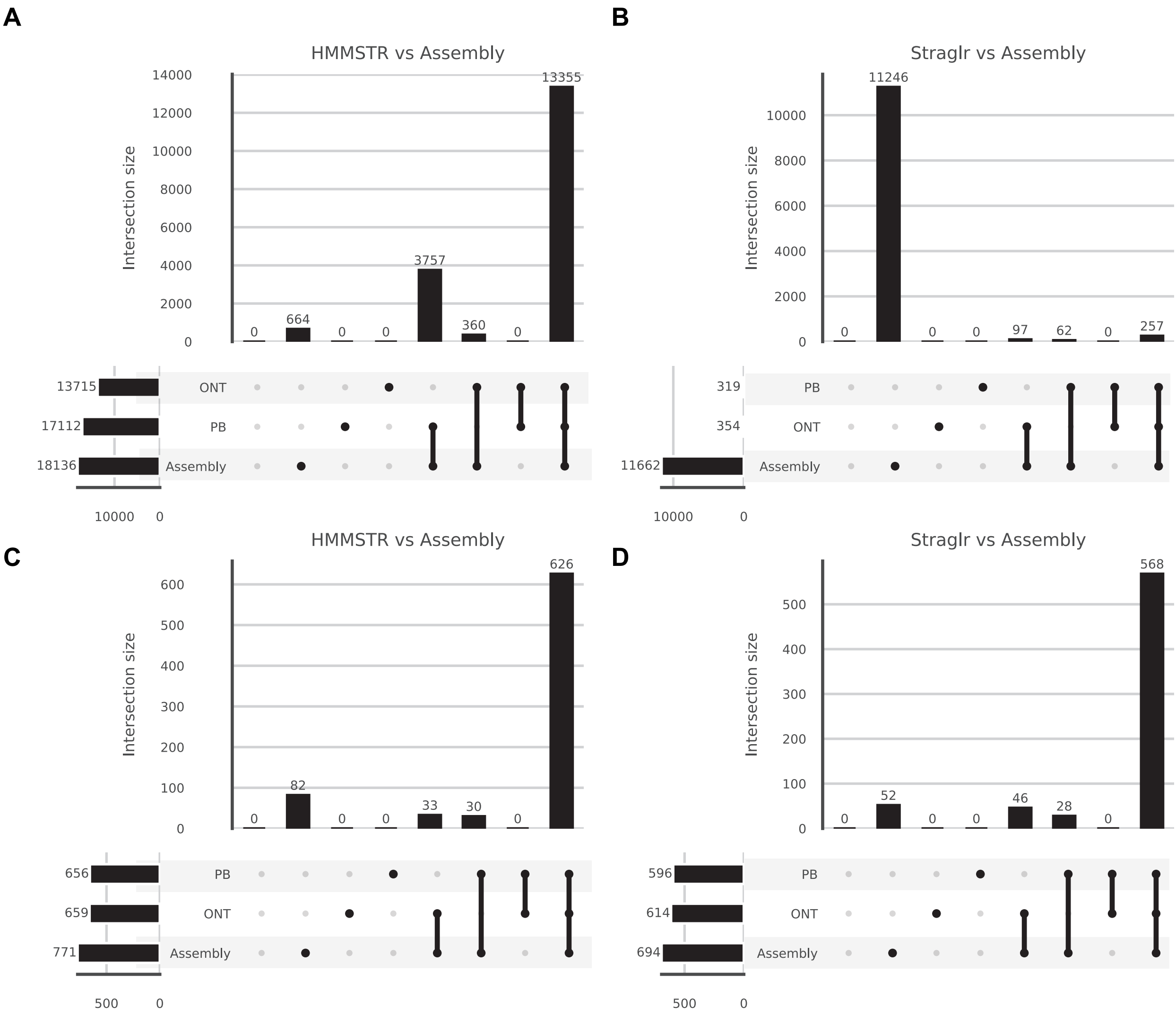


**Supplemental Figure 4: Heterozygous calls: over and under 100bp separation**

(**A** and **B)** Upset plots showing the number of regions called heterozygous with less than 100bp separation between alleles according to the GM12878 assembly across the ONT GM12878 dataset, PacBio CCS GM12878 dataset, and the HiFi assembly. (**A**) HMMSTR call results and (n=18,136) (**B**) Straglr call results (n=11,662) (**C** and **D**) Upset plots showing the number of regions called heterozygous with more than 100bp separation between alleles according to the GM12878 assembly across the ONT GM12878 dataset, a PacBio CCS GM12878 dataset, and the HiFi assembly. (**C**) HMMSTR call results and (n=771) (**D**) Straglr call results (n=694).

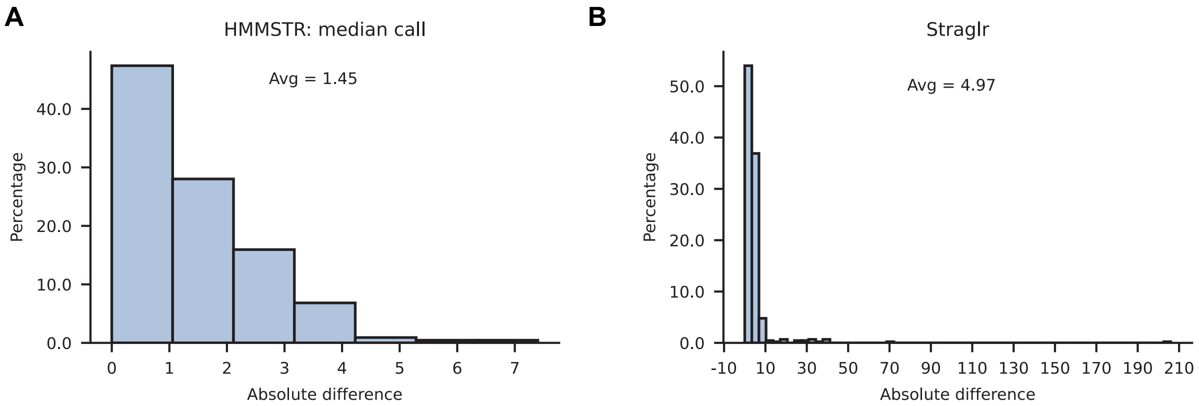


**Supplemental Figure 5: CHM13 STR Average Absolute Difference**

Average absolute difference between 439 long STR loci in CHM13 from nanopore sequencing of CHM13. (**A**) HMMSTR median calls. (**B**) Straglr calls.

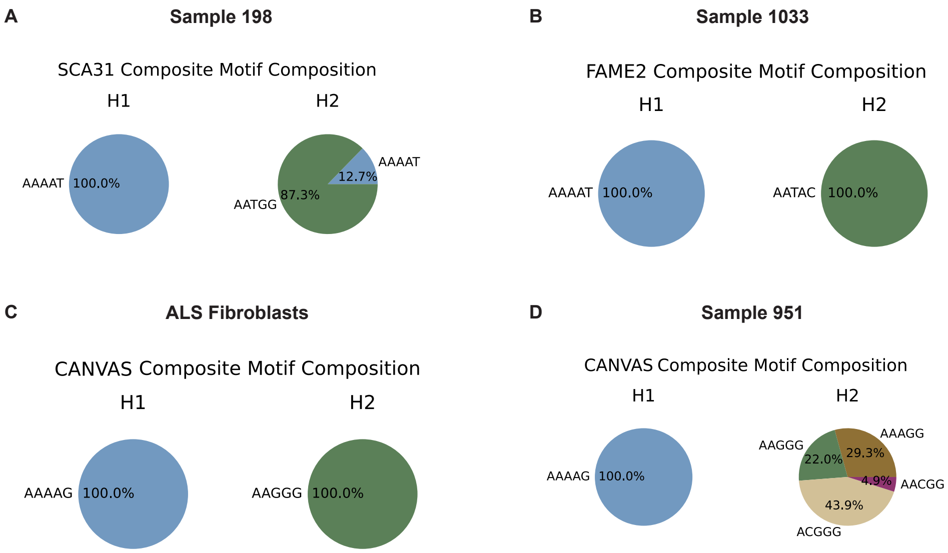


**Supplemental Figure 6: Motif composition at disease-associated loci in ALS/FTD samples**

Motif composition at tandem repeat loci in individuals with a neurodegenerative disease. Percentages are defined as the median number of motif occurrences across all reads in a given allele divided by the total median occurrences of all motifs found in the reads (**A**) Motif composition at expanded allele at *BEAN1* tandem repeat locus in individual 198, they carry one allele with inconsistent basecalls comprised of reads with the nested pathogenic AATGG motif or pure AAAAT motif (**B**) Motif composition at expanded *STARD7* tandem repeat locus in individual 103, they carry one allele with a motif with unknown disease-association, AATAC. (**C**) Motif composition at heterozygous pathogenic expansion (AAGGG) in *RFC1* in ALS fibroblasts. (**D**) Motif composition at rare allele at the *RFC1* locus associated with CANVAS in individual 951.

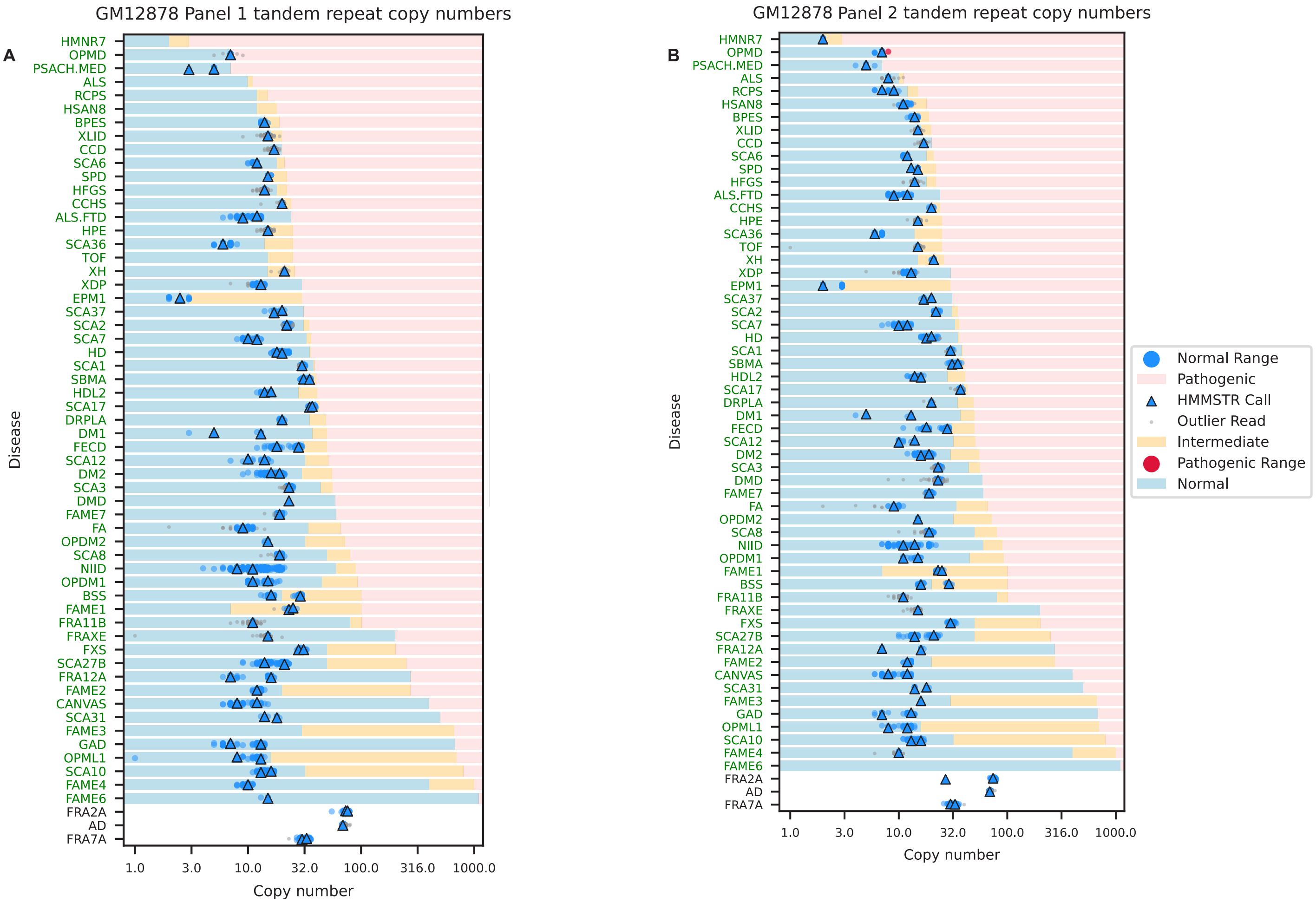


**Supplemental Figure 7: The swimlane plots show genotypes at disease associated target loci in GM12878**

The swimlane plots show genotypes across 54 (panel 1) or 60 (panels 2 and 3) disease associated target loci. All markers and ranges are displayed in log10 scale while the x-axis reflects the underlying repeat copy number. Dots indicate repeat counts for each read and triangles show HMMSTR median calls, where blue markings correspond to normal allele sizes and red indicate pathogenic length reads and calls. Gray dots indicate outlier copy number calls. Stars show fragmented reads that didn’t span the repeat (soft clipped in alignment). For each disease associated locus, the log copy number x-axis has been shaded to show the ranges of normal, intermediate, and pathogenic repeat copy numbers. For rows without shading, the ranges of normal and pathogenic lengths have not been described. Disease abbreviations (y-axis) shown in green indicate HMMSTR call was in the normal range for both alleles, purple indicates one allele was in the pathogenic range, and black indicates missing data or no ranges available. (**A**) Panel 1 GM12878 genotypes. (**B**) Panel 2 GM12878 genotypes.

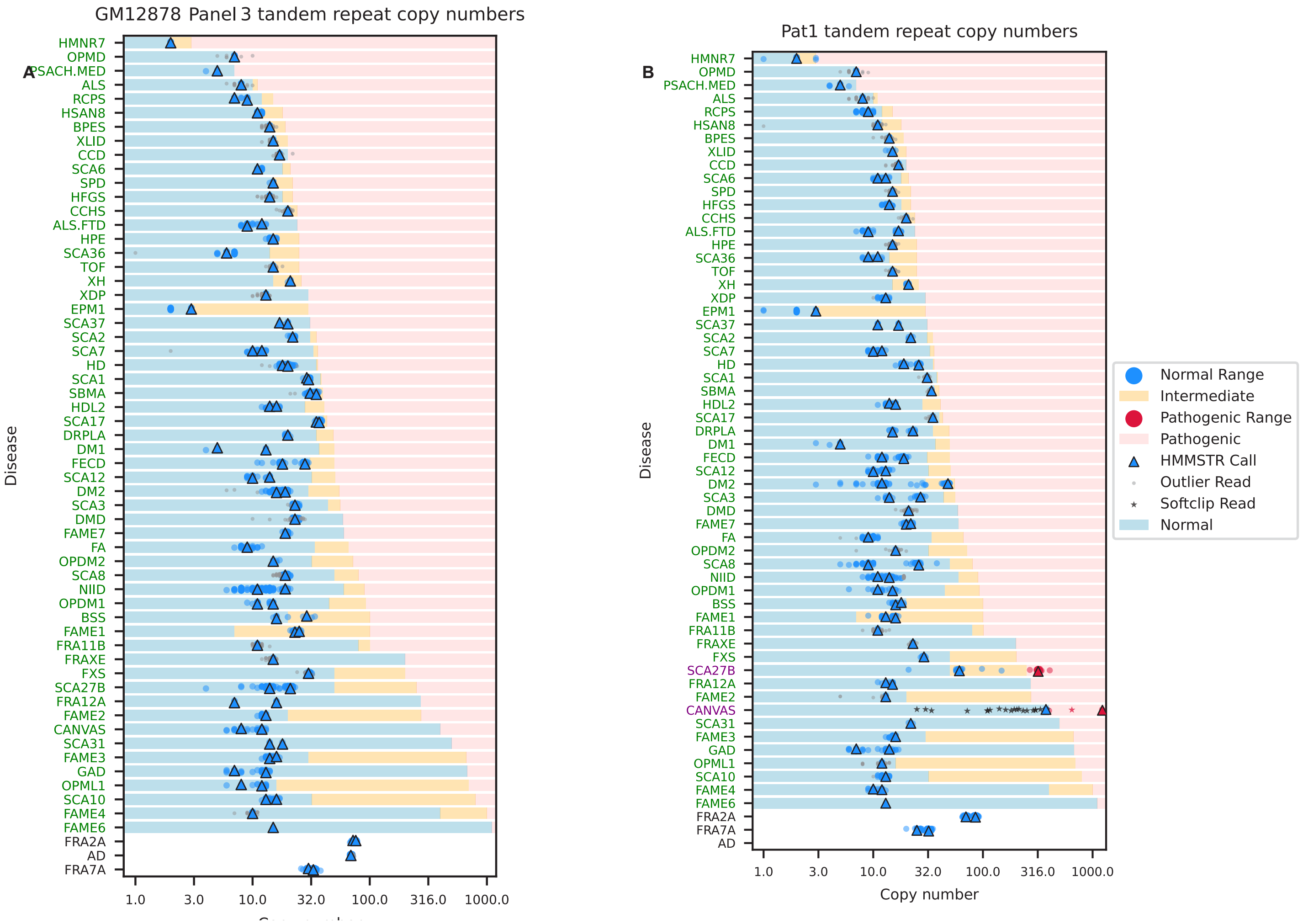


**Supplemental Figure 8: The swimlane plots show genotypes at disease associated target loci using Panel 3**

The swimlane plots show genotypes across 60 disease associated target loci. All markers and ranges are displayed in log10 scale while the x-axis reflects the underlying repeat copy number. Dots indicate repeat counts for each read and triangles show HMMSTR median calls, where blue markings correspond to normal allele sizes and red indicate pathogenic length reads and calls. Gray dots indicate outlier copy number calls. Stars show fragmented reads that didn't span the repeat (soft clipped in alignment). For each disease associated locus, the log copy number x-axis has been shaded to show the ranges of normal, intermediate, and pathogenic repeat copy numbers. For rows without shading, the ranges of normal and pathogenic lengths have not been described. Disease abbreviations (y-axis) shown in green indicate HMMSTR call was in the normal range for both alleles, purple indicates one allele was in the pathogenic range, and black indicates missing data or no ranges available. **(A)** Panel 3 GM12878 genotypes. **(B)** Panel 3 CANVAS PAT1 genotypes.

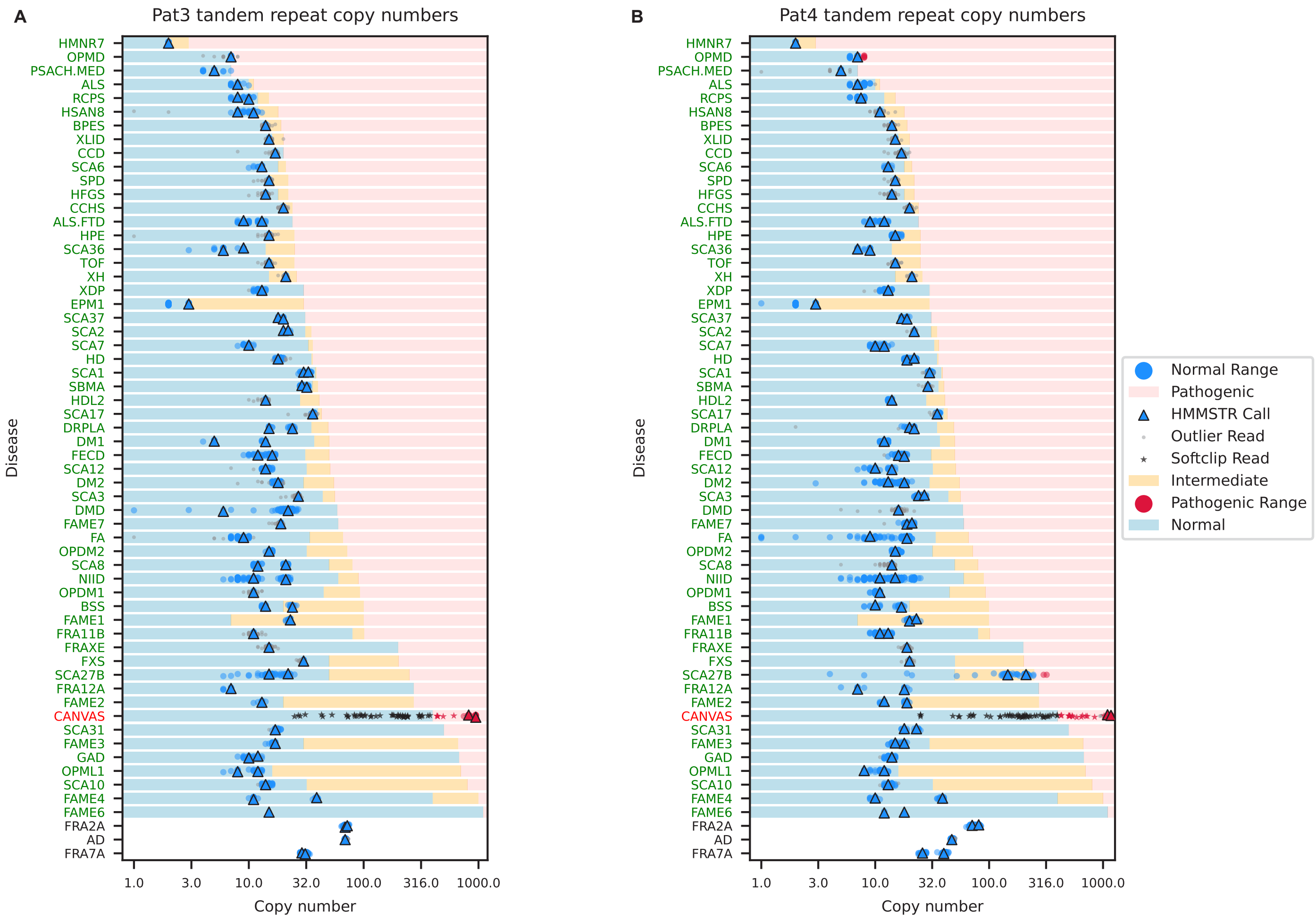


**Supplemental Figure 9: The swimlane plots show genotypes at disease associated target loci in two CANVAS patient-derived iPSC lines**

The swimlane plots show genotypes across 54 disease associated target loci. All markers and ranges are displayed in log10 scale while the x-axis reflects the underlying repeat copy number. Dots indicate repeat counts for each read and triangles show HMMSTR median calls, where blue markings correspond to normal allele sizes and red indicate pathogenic length reads and calls. Gray dots indicate outlier copy number calls. Stars show fragmented reads that didn't span the repeat (soft clipped in alignment). For each disease associated locus, the log copy number x-axis has been shaded to show the ranges of normal, intermediate, and pathogenic repeat copy numbers. For rows without shading, the ranges of normal and pathogenic lengths have not been described. Disease abbreviations (y-axis) shown in green indicate HMMSTR call was in the normal range for both alleles, purple indicates one allele was in the pathogenic range, red indicates a biallelic expansion, and black indicates missing data or no ranges available. (**A**) Panel 1 Pat3 genotypes. (**B**) Panel 1 Pat4 genotypes.

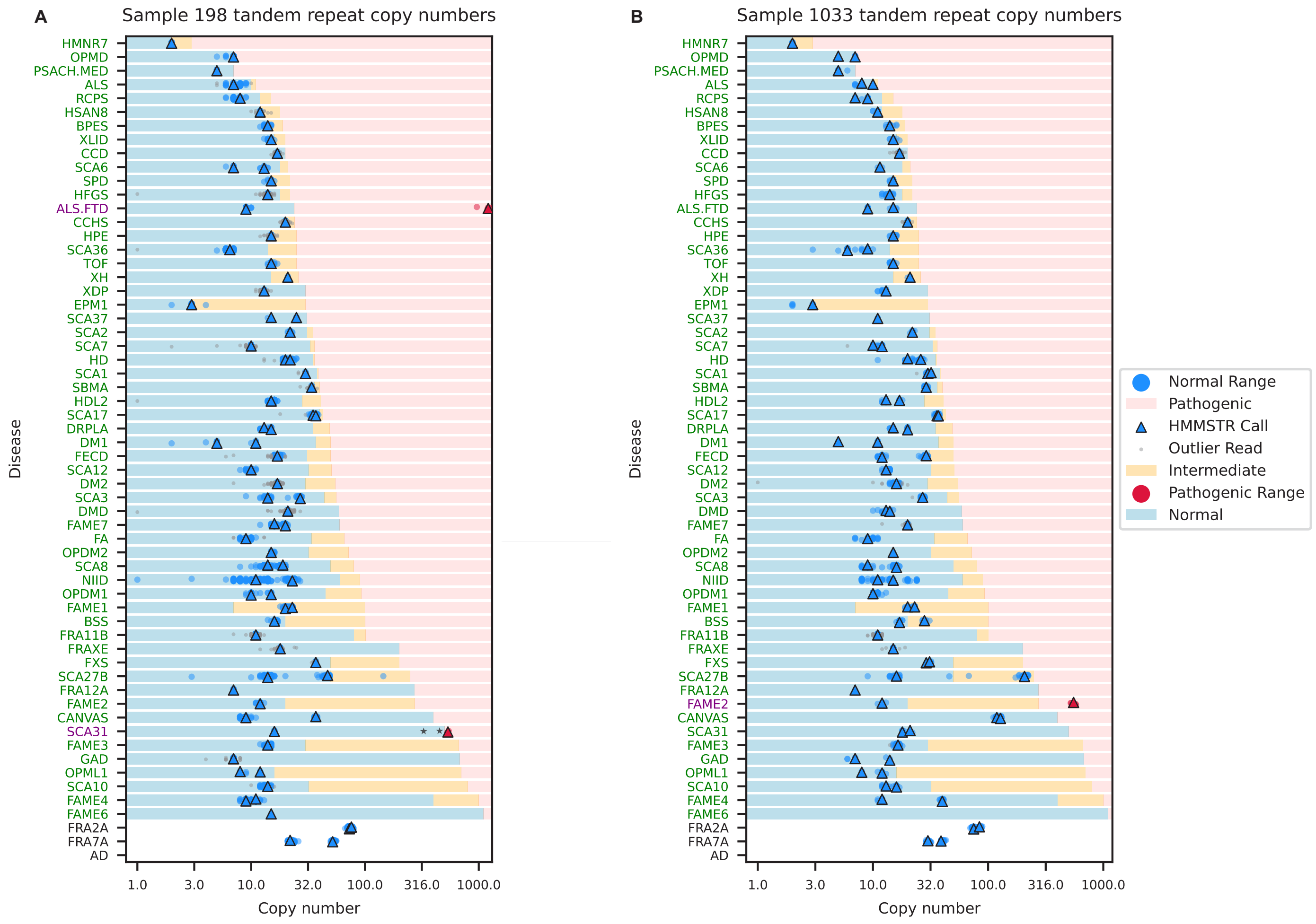


**Supplemental Figure 10: The swimlane plots show genotypes at disease associated target loci in ALS/FTD samples**

The swimlane plots show genotypes across 54 (panel 1) or 60 (panels 2 and 3) disease associated target loci. All markers and ranges are displayed in log10 scale while the x-axis reflects the underlying repeat copy number. Dots indicate repeat counts for each read and triangles show HMMSTR median calls, where blue markings correspond to normal allele sizes and red indicate pathogenic length reads and calls. Gray dots indicate outlier copy number calls. Stars show fragmented reads that didn't span the repeat (soft clipped in alignment). For each disease associated locus, the log copy number x-axis has been shaded to show the ranges of normal, intermediate, and pathogenic repeat copy numbers. For rows without shading, the ranges of normal and pathogenic lengths have not been described. Disease abbreviations (y-axis) shown in green indicate HMMSTR call was in the normal range for both alleles, purple indicates one allele was in the pathogenic range, and black indicates missing data or no ranges available. (**A**) Panel 3 genotypes for individual 198. (**B**) Individual 1033 genotypes using Panel 3.

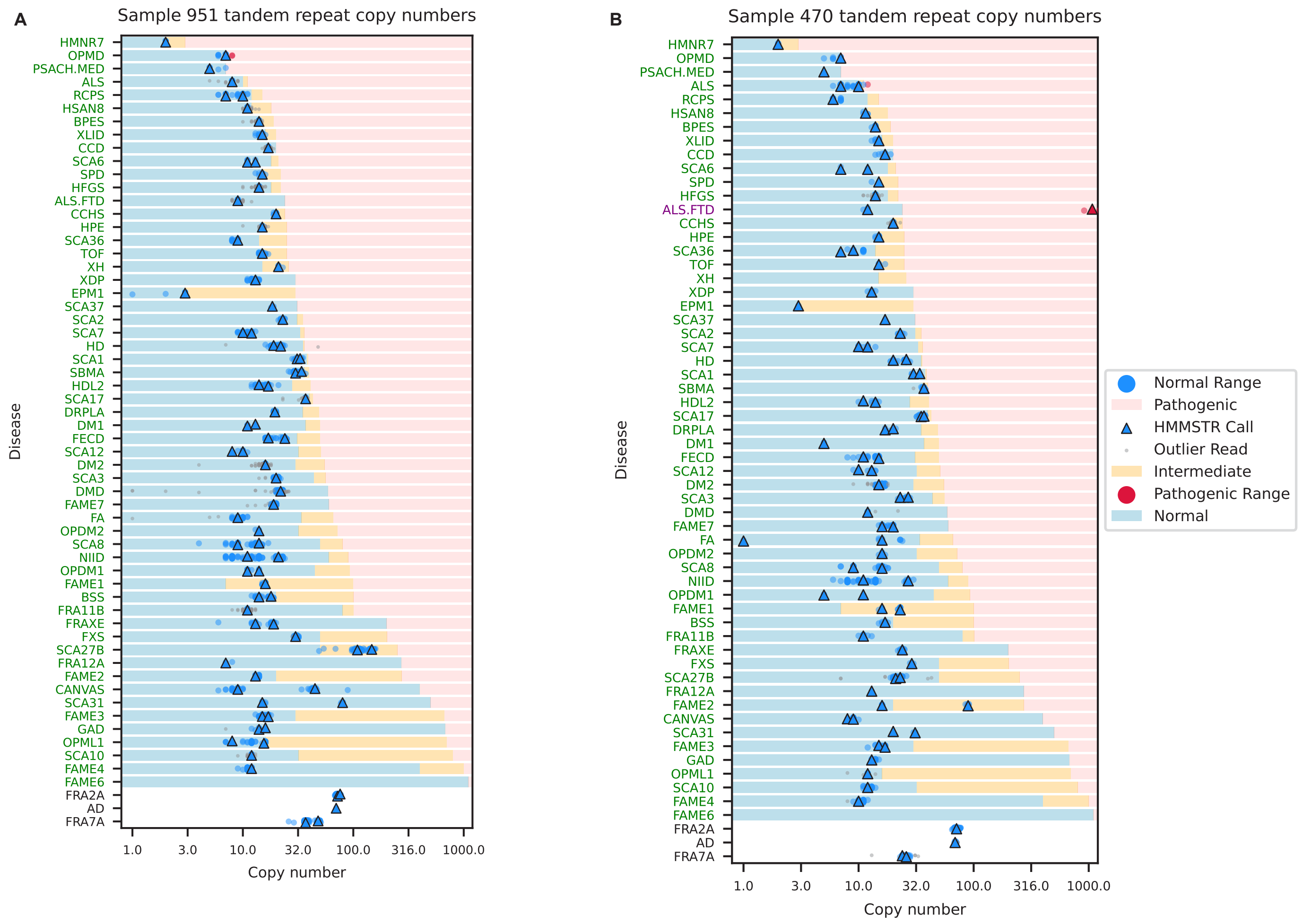


**Supplemental Figure 11: The swimlane plots show genotypes at disease associated target loci in ALS/FTD samples**

The swimlane plots show genotypes across 60 disease associated target loci. All markers and ranges are displayed in log10 scale while the x-axis reflects the underlying repeat copy number. Dots indicate repeat counts for each read and triangles show HMMSTR median calls, where blue markings correspond to normal allele sizes and red indicate pathogenic length reads and calls. Gray dots indicate outlier copy number calls. Stars show fragmented reads that didn’t span the repeat (soft clipped in alignment). For each disease associated locus, the log copy number x-axis has been shaded to show the ranges of normal, intermediate, and pathogenic repeat copy numbers. For rows without shading, the ranges of normal and pathogenic lengths have not been described. Disease abbreviations (y-axis) shown in green indicate HMMSTR call was in the normal range for both alleles, purple indicates one allele was in the pathogenic range, and black indicates missing data or no ranges available. (**A**) Individual 951 genotypes using Panel 3. (**B**) Individual 470 genotypes using Panel 3.

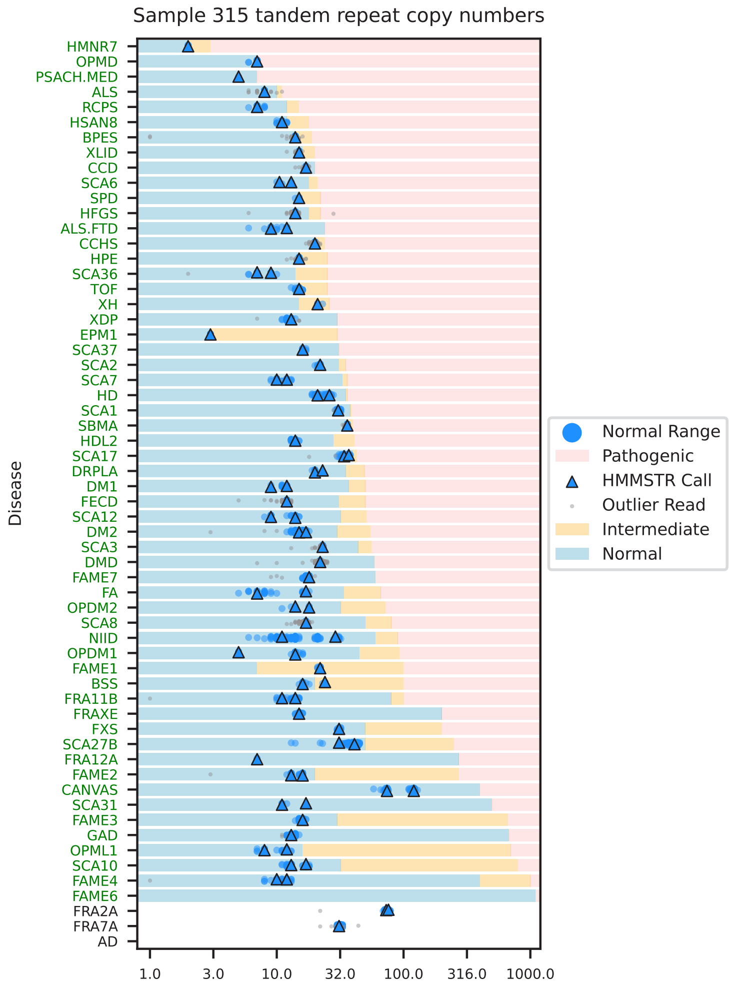


**Supplemental Figure 12: The swimlane plots show genotypes at disease associated target loci in ALS/FTD samples**

The swimlane plots show genotypes across 60 disease associated target loci. All markers and ranges are displayed in log10 scale while the x-axis reflects the underlying repeat copy number. Dots indicate repeat counts for each read and triangles show HMMSTR median calls, where blue markings correspond to normal allele sizes and red indicate pathogenic length reads and calls. Gray dots indicate outlier copy number calls. Stars show fragmented reads that didn't span the repeat (soft clipped in alignment). For each disease associated locus, the log copy number x-axis has been shaded to show the ranges of normal, intermediate, and pathogenic repeat copy numbers. For rows without shading, the ranges of normal and pathogenic lengths have not been described. Disease abbreviations (y-axis) shown in green indicate HMMSTR call was in the normal range for both alleles, purple indicates one allele was in the pathogenic range, and black indicates missing data or no ranges available. Individual 315 genotypes using Panel 3.

**A**

| Dataset | Dataset Coverage | Target Coverage | # of Regions Genotyped: HMMSTR | # of Regions Genotyped: Straglr | # of Regions Genotyped: Shared After Filter |
|---|---|---|---|---|---|
| ONT GM12878 | 34.9 | 21.4 | 9,486 | 8,687 | 3,305 |
| PacBio CCS | 27.6 | 31.0 | 11,007 | 10,967 | 4,407 |

**B**

| Dataset | Dataset Coverage | Target Coverage | # of Regions Genotyped: HMMSTR | # of Regions Genotyped: Straglr | # of Regions Genotyped: Shared After Filter |
|---|---|---|---|---|---|
| ONT GM12878 | 34.9 | 41.1 | 18,938 | 12,885 | 5,737 |
| PacBio CCS | 27.6 | 30.4 | 18,911 | 12,363 | 5,460 |

**Supplemental Figure 13: Summary of dataset coverage and regions genotyped**

Summary of overall coverage and coverage over target loci both ONT and PacBio CCS datasets as well as number of regions successfully genotyped and used in benchmarking analysis for (**A**) Figure 1 benchmark and (**B**) Figure 2 (heterozygous) benchmark.

| |
|---|
| **Supplementary Table 1:** Table containing plasmid constructs used for HMMSTR development and benchmarking including motifs, PCR estimated copy numbers, and the restriction enzymes used for sequencing. |
| **Supplementary Table 2:** Panel 1. Table lists the disease associated tandem repeat targets in Panel 1, including the target coordinates, gene, disease and abbreviation. The repeat coordinates for *FAME3* and *FAME6* are incorrect in this list. |
| **Supplementary Table 3:** Panel 1 guides. List of the 118bp oligos used in Panel 1. |
| **Supplementary Table 4:** Panel 2. Table lists the disease associated tandem repeat targets in Panel 2, including the target coordinates, gene, disease and abbreviation. The repeat coordinates for *FAME3* and *FAME6* are incorrect in this list. |
| **Supplementary Table 5:** Panel 2 guides. List of the 118bp oligos used in Panel 2. |
| **Supplementary Table 6:** Panel 3. Table lists the disease associated tandem repeat targets in Panel 3, including the target coordinates, gene, disease and abbreviation. The repeat coordinates for *FAME6* are incorrect in this list. |
| **Supplementary Table 7:** Panel 3 guides. List of the 118bp oligos used in Panel 3. |
| **Supplementary Table 8:** Coverage results from GM12878 nCATS targeted sequencing across rounds of panel optimization. |
